# Neurocomputational evidence of sustained Self-Other mergence after psychedelics

**DOI:** 10.1101/2025.10.07.25337510

**Authors:** Pablo Mallaroni, Natasha L. Mason, Katrin H. Preller, Adeel Razi, Sam Ereira, Johannes G. Ramaekers

## Abstract

Mental illness is often characterised by a maladaptive sense of *self*. The neurobiological basis of Self-Other distinction may provide targets for therapeutic interventions. Psychedelics alter the experience of selfhood, but the neurocomputational mechanism is unclear. We used a computationally-informed behavioural assay to investigate whether psychedelics disrupt Self-Other boundaries in belief formation. In a double-blind, crossover design, 22 participants received placebo, psilocybin or 2C-B (2,5-dimethoxy-4-bromophenethylamine). The next day, we fitted reinforcement learning models to probabilistic false-belief task behaviour, yielding objective Self-Other distinction measures. Compared to placebo, psychedelics induced a state of Self-Other mergence, associated with a multivariate signal of sustained psychosocial wellbeing. Effective-connectivity estimates from resting-state fMRI showed that this behavioural change was associated with reduced inhibitory tone from right temporoparietal junction to dorsomedial prefrontal cortex. We show that psychedelics quantifiably act on the neural basis of Self-Other distinction, offering potential routes to precision therapeutics in psychedelic psychiatry.

## Introduction

Humans have evolved the ability to represent the contents of each others’ minds. This ability, to *put myself into your shoes*, is often referred to as Theory of Mind^1,2^ or mentalising^3^. It allows humans to make predictions about each other^4,5^, and may be a fundamental process behind the emergence of human social networks and societies^6,7^.

A critical component of Theory of Mind is the selective attribution of a mental state, such as a belief or a desire, to a specific agent, such as Self or Other. Neuroimaging and electrophysiological research show that humans can achieve this by simulating the neural computations of other people in segregated, agent-specific, neural circuits^8–12^. However, Self-Other distinction is a flexible process that adapts to different social environments^11^. On one hand, it would be unhelpful for an individual to persistently conflate the contents of their own mind with the contents of another person’s mind^13^. On the other hand, empathic responding, where another person’s mental states are experienced as one’s own, can be adaptive to facilitate social bonding and mutual understanding^14–19^.

As well as enabling complex social behaviour, adaptive Self-Other distinction is necessary for furnishing us with a coherent sense of Self^20,21^. It is therefore unsurprising that aberrant mentalising is a transdiagnostic feature of several mental illnesses^10,22^, including autism spectrum disorder (ASD)^23,24^, schizophrenia^25,26^, borderline personality disorder (BPD)^27,28^, depression^29^ and dementia^30^. People with these conditions experience difficulties in maintaining social connections and a coherent sense of Self, highlighting the potential value in identifying interventions that selectively modulate the neurobiological basis of Self-Other distinction.

Serotonergic psychedelics are one class of drugs that can transiently and profoundly alter a person’s experience of selfhood. Compounds such as psilocybin, lysergic acid diethylamide (LSD), mescaline, 2,5-dimethoxy-4-bromophenethylamine (2C-B), and dimethyltryptamine (DMT) reliably induce a blurring of the boundary between Self and non-Self^31–33^, likely via agonism at 5-HT_2A_ receptors^34,35^. This acute state, known as *ego dissolution*^36–38^, is thought to underpin the observed therapeutic effects of psychedelics in disorders marked by aberrant Self-referential processing^39–42^. In the days following the acute phase, there is often a subacute *afterglow* period of sustained psychosocial wellbeing, characterised by enhanced empathy, suggestibility, and a sense of social connectedness^43–48^.

The neurocomputational basis of ego dissolution and the ensuing afterglow period are not well understood^49^. Experimental work on Self-Other processing under psychedelics has largely relied on subjective reports. Where behaviour has been assessed, studies consistently show that psychedelics induce a blurring of the Self-Other boundaries of affective and sensory states^50–52^. However, there has been little work to generate behavioural data amenable to computational modelling. Computationally-informed behavioural tasks, such as that used in the current study, elicit behavioural data that can test explicit models of underlying cognitive processes. By moving from subjective reports to objective computational metrics, it may be possible to describe and explain psychedelic alterations in Self-Other processing within established neurocomputational frameworks.

Here within a double-blind, placebo-controlled, within-subject crossover design, we tested whether the psychedelic compounds, psilocybin and 2C-B, could induce enduring changes in Self-Other distinction with respect to prediction errors, fundamental computational units in learning and belief formation^53,54^. We employed a probabilistic false-belief task (pFBT) to record behavioural data and estimate subject-specific behavioural parameters. These parameters provide an objective measurement of a person’s propensity to process their environment using agent-specific prediction errors or agent-independent prediction errors^10,11,27^. Agent-specific prediction errors are sensory surprise signals that contain information not only about the magnitude of the surprise, but also about the identity of the person who is experiencing the surprise (e.g. *me* or *you*), allowing for Self-Other distinction in belief states. Agent-independent prediction errors encode the surprise signal without any information about agent identity, allowing for a Self-Other mergence in belief formation. The pFBT takes inspiration from the field of computational psychiatry^22,55–57^, which seeks to identify objective biomarkers, by explaining cognition and mental illness using mathematical models. These models are constrained by quantitative parameters, estimated from a person’s behavioural, neural or other physiological data.

To complement these behavioural measures, participants were scanned with ultra high field 7 Tesla resting-state fMRI (rsfMRI) during peak drug effects. By estimating effective connectivity using spectral dynamic causal modelling (DCM), we aimed to identify acute drug-induced connectivity changes, between brain regions known to be involved in Self-Other processing, that were associated with behavioural effects.

By employing tools from computational psychiatry, our aim was to assess whether and how psychedelic compounds alter an objective measure of Self-Other distinction in learning and belief formation. We predicted that psychedelics shift people into a cognitive state where they were more likely to conflate the mental states of Self and Other, and that this altered state is associated with the broad psychosocial wellbeing previously observed in people treated with psychedelics.

## Results

### Psychedelics do not impair performance on the probabilistic false belief task

N = 22 participants were treated with matched doses of 15mg psilocybin, 20mg 2C-B, and placebo, in a randomised order, with two weeks between each dosing session (Fig. 1a). One day after each dosing session, we administered a probabilistic false belief task (pFBT)^10^, in order to measure components of Self-Other processing in the subacute phase following psychedelic treatment.

**Figure 1.**
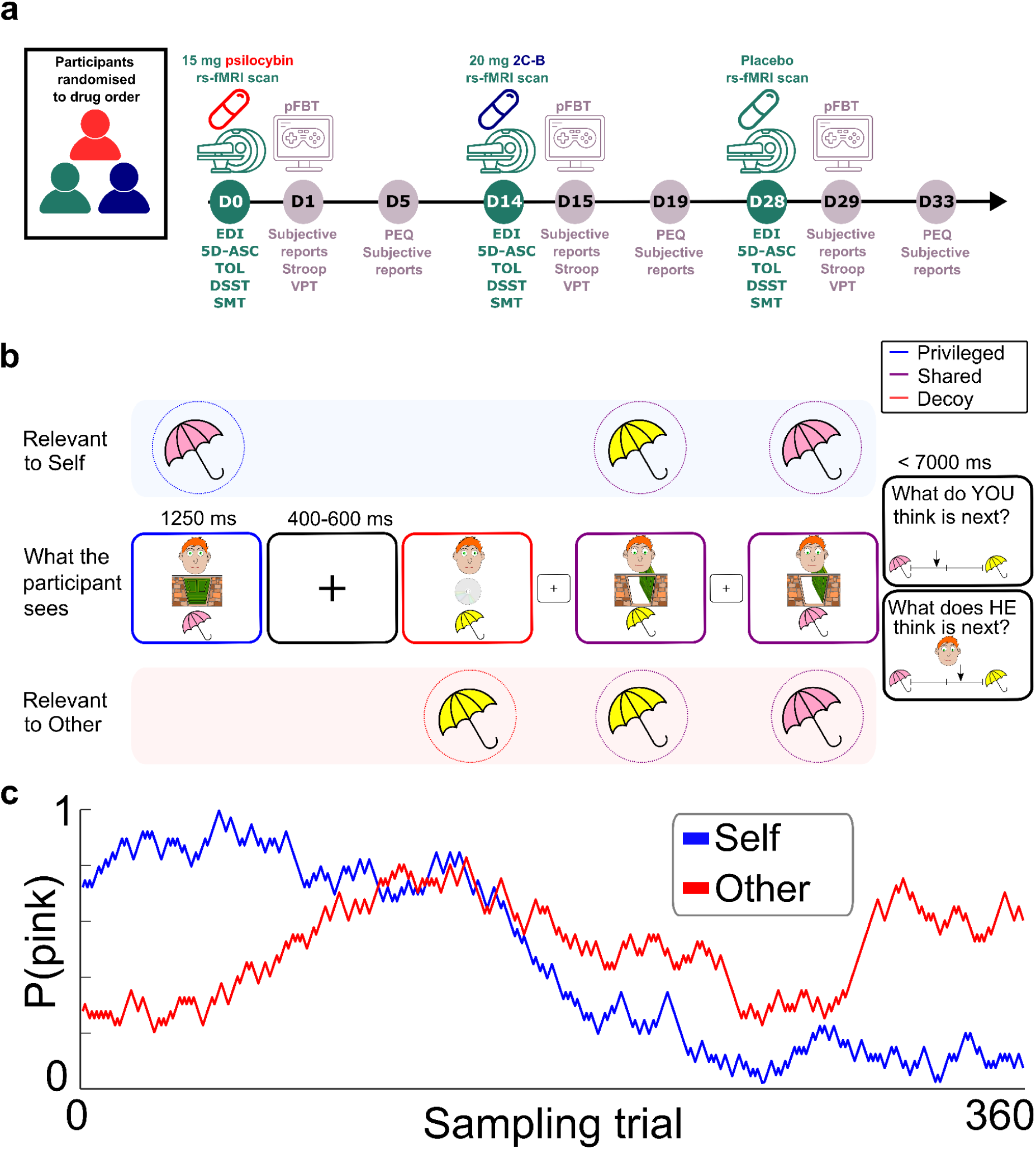
- Experimental design. **a**, This was a randomised crossover study where participants received psilocybin, 2C-B and placebo, in a randomised order, with a 2 week washout between each dosing day. On each dosing day (days 0, 14 and 28) rsfMRI data was collected and participants were assessed using the Ego Dissolution Inventory (EDI) and 5-Dimensions of Altered States of Consciousness (5D-ASC) questionnaire. They also completed several cognitive tasks: Tower of London (TOL), Digit Symbol Substitution Test (DSST) and a Spatial Memory Test (SMT). The day after each dosing day (days 1, 15 and 29), participants completed a probabilistic false belief task (pFBT), a Stroop task and a visuospatial perspective-taking task (VPT). The following subjective self-report questionnaires were administered: Inclusion of Other in the Self (IOS), Positive and Negative Affect Schedule (PANAS), Social Connectedness Scale–Revised (SCS-R). Five days after each dosing day (days 5, 19 and 33) the same battery of questionnaires was administered, in addition to the persisting effects questionnaire (PEQ). **b**, pFBT task design. Participants observed a series of *sampling trials* where each trial revealed either a pink umbrella (with probability *P*) or a yellow sun-shade (with probability 1-*P*). Participants were required to infer and keep track of *P* as well as another agent’s false belief about *P.* On *privileged* trials the information was hidden from the other agent. On *shared* trials, the information was available to the other agent. On *decoy* trials, the information was available to the other agent, but unbeknown to the other agent, the information was misleading. Participants were intermittently probed to report their estimate of *P* or the estimate of the other agent’s belief about *P*. **c**, The pFBT was designed such that the drifting variable *P* (blue) and the other agent’s belief about *P* (red) were not correlated.

The pFBT (Fig. 1b) presented participants with a sequence of binary samples, in this case yellow sun-shades or pink umbrellas. Each sample was paired with a second image indicating whether or not a fictional other person could see the sample. On some trials, this image indicated that the information *could* be seen by the other person, but that it was misleading. The probability of a trial revealing a pink umbrella gradually drifted throughout the task along a random walk (Fig. 1c, blue line). Participants were instructed to infer and keep track of this drifting probability so that they could make accurate predictions about what the outcome might be on the next trial. By combining a randomised mixture of trials where the other agent could see some outcomes and not others, and also saw some misleading outcomes, the other person’s belief about the probability also followed a random walk, decorrelated from the true underlying probability (Fig. 1c, red line). Participants were instructed to also infer and keep track of the other person’s false belief about the probability. Behavioural data was collected on probe trials, in which participants reported their estimates of the probability, or their estimates of the other person’s false belief about the probability.

Accuracy on the pFBT was calculated as the correlation between participants’ reports on probe trials, and the true underlying probabilities generating outcomes relevant to Self and Other (see Methods). Most participants performed better than chance, in all three testing sessions (Fig. 2a), demonstrating a good understanding of the task. However, two participants performed below chance-level in multiple sessions, and they were excluded from all further analyses.

**Figure 2.**
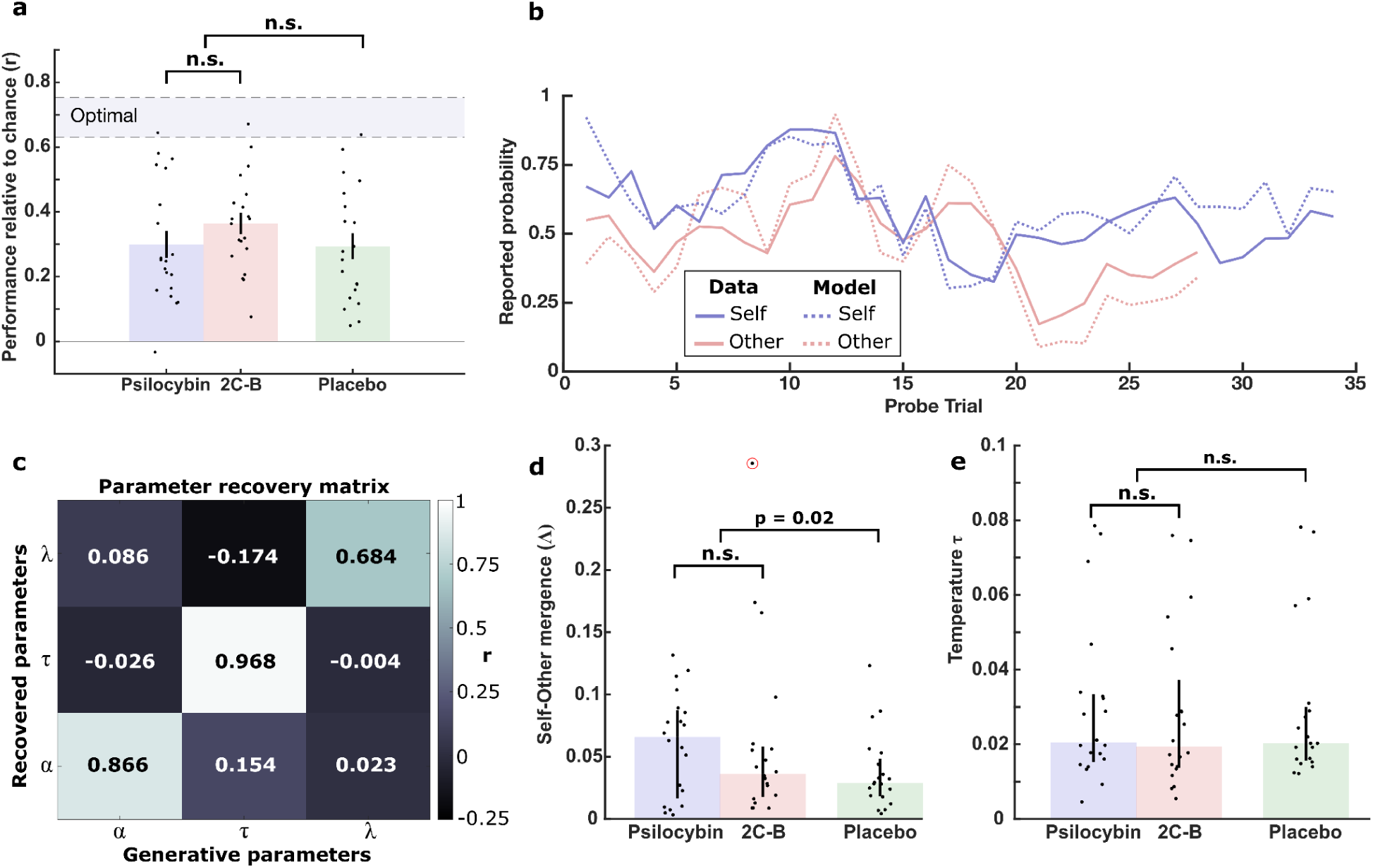
- Analysis of pFBT behavioural task data. **a**, Task performance, calculated as the correlation between participant reports and true underlying probabilities generating *sampling trials*. As described in Methods, “optimal” performance was estimated by simulating observers with perfect memory and no behavioural stochasticity. There was no difference in performance between post-2C-B sessions and post-psilocybin sessions. There was also no difference in performance between post-placebo sessions and post-psychedelic sessions. The bar graph shows the median +/- interquartile range (IQR) and each dot represents a different participant. N = 20. **b**, Exemplar model fit for a single participant session. To avoid cherry-picking, the participant session with the median log-model evidence across all participant sessions was selected for visualisation. Model predictions are illustrated for *Self probe* and *Other probe* trials separately. **c**, Parameter recovery matrix. Pearson correlation coefficients between generative parameters of 1000 simulated participants and recovered parameters demonstrate that the three model parameters (α/learning rate, τ /temperature, λ/leak) are separately identifiable. **d**, Self-Other mergence (Λ, i.e. product of α and |λ|) was significantly lower following placebo administration than following psychedelic administration. The bar graph shows the median +/- IQR and each dot represents a different participant. Also see Figure S1b for difference scores between drug conditions and placebo. One datapoint for the 2C-B session, circled in red, was identified as an outlier. In a sensitivity analysis where this participant was excluded, statistical results were consistent. N = 20. **e**, There was no difference in τ/temperature between post-placebo and post-psychedelic sessions. The bar graph shows the median +/- IQR and each dot represents a different participant. N = 20.

For the remaining N = 20 participants, we fitted a linear mixed-effects model, predicting accuracy from condition (placebo, psilocybin or 2C-B) with random intercepts estimated for each participant, to account for repeated measures. Overall, accuracy was significantly higher than chance (*F*_1,57_ = 60.97, *p* < 0.001) and there was no effect of condition on accuracy (*F*_2,57_ = 1.69, *p* = 0.19) These results suggest that during the subacute phase following psychedelic administration, participants’ comprehension of and ability to perform the pFBT was not impaired.

### Psychedelics promote Self-Other mergence in prediction errors

We fitted a family of reinforcement-learning style models to participants’ behavioural data from probe trials (see Methods). These models shared the assumption that, on each trial, participants updated their own belief about the underlying probability, and also updated a simulation of the other agent’s belief using a parallel and simultaneous belief-update. This family of models has been shown to approximate behaviour in the pFBT well^10,11,27^ and also furnish computational variables that can explain brain activity during task engagement^10,11^.

The models were nested, and they differed in whether they allowed the parallel belief-updates for Self and Other to be parameterised independently or jointly, and whether or not they allowed for cross-agent leakage. In one version of the model, the parallel belief-updates were entirely segregated, reflecting a perfect implementation of Self-Other distinction. In another version of the model, the participant’s prediction errors were leaky, leading them to erroneously update their simulation of the other agent’s belief. This *idiocentric updating* ^27^, reflects a situation where *my own beliefs influence my simulation of your beliefs*. In an alternative version of the model, the simulated prediction errors of the other agent were leaky, leading to erroneous updating of the participant’s own beliefs. This *allocentric updating* ^27^, reflects a situation where *my simulation of your beliefs influences my own beliefs*.

In a Bayesian model comparison of 36 models (see Methods) we found that the best model to explain behaviour across all three testing sessions was a three parameter model. The model specifies a learning rate (α), which governs the rate at which new information is incorporated into old beliefs (i.e. sensitivity to surprise), a decision temperature (τ), which governs behavioural stochasticity in decision-making, and a Self-Other leak parameter (λ), governing bi-drectional leakage from Self-to-Other and Other-to-Self, capturing both idiocentric and allocentric updating. This model explained true choice data well (Fig. 2b) with r^2^ = 0.41 ± 0.24 (median ± IQR). The three parameters were recoverable and separately identifiable (Fig. 2c).

Previous studies using the pFBT have found evidence for interindividual variability and context-dependent effects in whether subjects make use of idiocentric leak, allocentric leak, neither, or both, in belief-updating ^11,27^. A model recovery analysis (Fig. S1d) showed excellent model recovery, indicating that the task, dataset and model fitting procedure of the current study were adequately powered and robust to discriminate different specifications of leak parameter λ. Unlike the current study, the most recent published work using the pFBT found that group data were best explained with a model with two leak parameters for independent idiocentric and allocentric updating; that previous study investigated a clinical cohort of people with borderline personality disorder, who showed differences from matched controls specifically in idiocentric leakage^27^. Parameter values in the current dataset (Fig S1a) were comparable to those estimated in previous datasets.

We were specifically interested in whether psychedelic compounds alter the sensitivity to leaked prediction errors. As described in Methods, this is captured by the parameter Λ, which is the product of α and |λ| and reflects the extent to which *my belief is influenced by my simulation of your prediction errors* and *my simulation of your belief is influenced by my own prediction errors*. In other words, Λ is a metric that quantifies Self-Other mergence in belief updating. We did not predict any differences between 2C-B and psilocybin. Consistent with this, there was no difference in Self-Other mergence (Λ) between the post-psilocybin session and the post-2C-B session (t(19) = –0.048, *p* = 0.96).

As two primary 5-HT_2A_ agonists, we had no a priori predictions about differences between 2C-B and psilocybin on Self-Other mergence. To capture the overall effect of either psychedelic compound, we fitted a linear mixed-effects model predicting Self-Other mergence (Λ) from whether the session involved drug or placebo (binary). The model included random intercepts and random condition-specific slopes for each participant, allowing the effect of each drug (placebo, 2C-B, psilocybin) to vary by subject (see Methods). A likelihood ratio test favoured this model compared to a nested model with only random intercepts (χ²(5) = 16.80, *p* = .0049). We found that psychedelics significantly increased Self-Other mergence relative to placebo (F_1,57_ = 5.77, *p* = 0.02). In a sensitivity analysis, we refitted the model after excluding one participant’s 2C-B session, which had an anomalously high Self-Other mergence metric (Fig. 2d); results were consistent (F_1,57_ = 4.65, *p* = 0.035). Drug-specific contrasts were significant for psilocybin (F_1,56_ = 4.98, *p* = 0.029, *d* = 0.49) but not 2C-B (F_1,56_= 1.63, *p* = 0.21, *d* = 0.31). The two drug effects did not differ from each other (F_1,56_ = 0.002, *p* = 0.9606) with comparable mean increases (psilocybin: Δ = 0.021; 2C-B: Δ = 0.024) and greater within-subject response variability under 2C-B (SD = 0.079 vs 0.043).

We noted in our results a dissociation between task performance and pFBT parameters; despite the fact that the psychedelic compounds promoted Self-Other mergence (Fig. 2d), task accuracy remained unimpaired (Fig. 2a). In a simulation based analysis (fig. S1c) we found that indeed these two metrics were only associated with each other when parameter values were extreme. When Self-Other mergence was constrained to the empirical distribution of our dataset, it was not associated with impaired task performance. As expected, task accuracy was negatively associated with the temperature parameter τ.

Fitting another linear mixed effects model predicting decision temperature (Fig. 2e) instead of Self-Other mergence, we found no effect of psychedelics on decision temperature (F_1,57_ = 0.54, *p* = 0.82). There was also no drug effect on learning rate (F_1,57_ = 0.54, *p* = 0.59), suggesting that it was not simply enhanced sensitivity to surprise that was driving the effect on Self-Other mergence (Λ). Taken together, the results suggest that these psychedelic compounds induce a specific effect on belief formation, wherein Self-Other mergence is enhanced in the subacute phase following drug administration,in the absence of cognitive impairment

### Psychedelic-induced Self-Other mergence predicts (sub)acute subjective effects

To assess whether drug administration was accompanied by changes in subjective changes in psychosocial wellbeing, we characterised acute ego dissolution using the Ego Dissolution Inventory (EDI) and the 5-Dimensions of Altered States of Consciousness (5D-ASC), and follow-up measures using the Persisting Effects Questionnaire (PEQ), Positive and Negative Affect Scale (PANAS), the Inclusion of Other in the Self (IOS) scale, and the Social Connectedness Scale–Revised (SCS-R) (see Fig. 3a, Methods and Supplementary Table 1).

**Figure 3.**
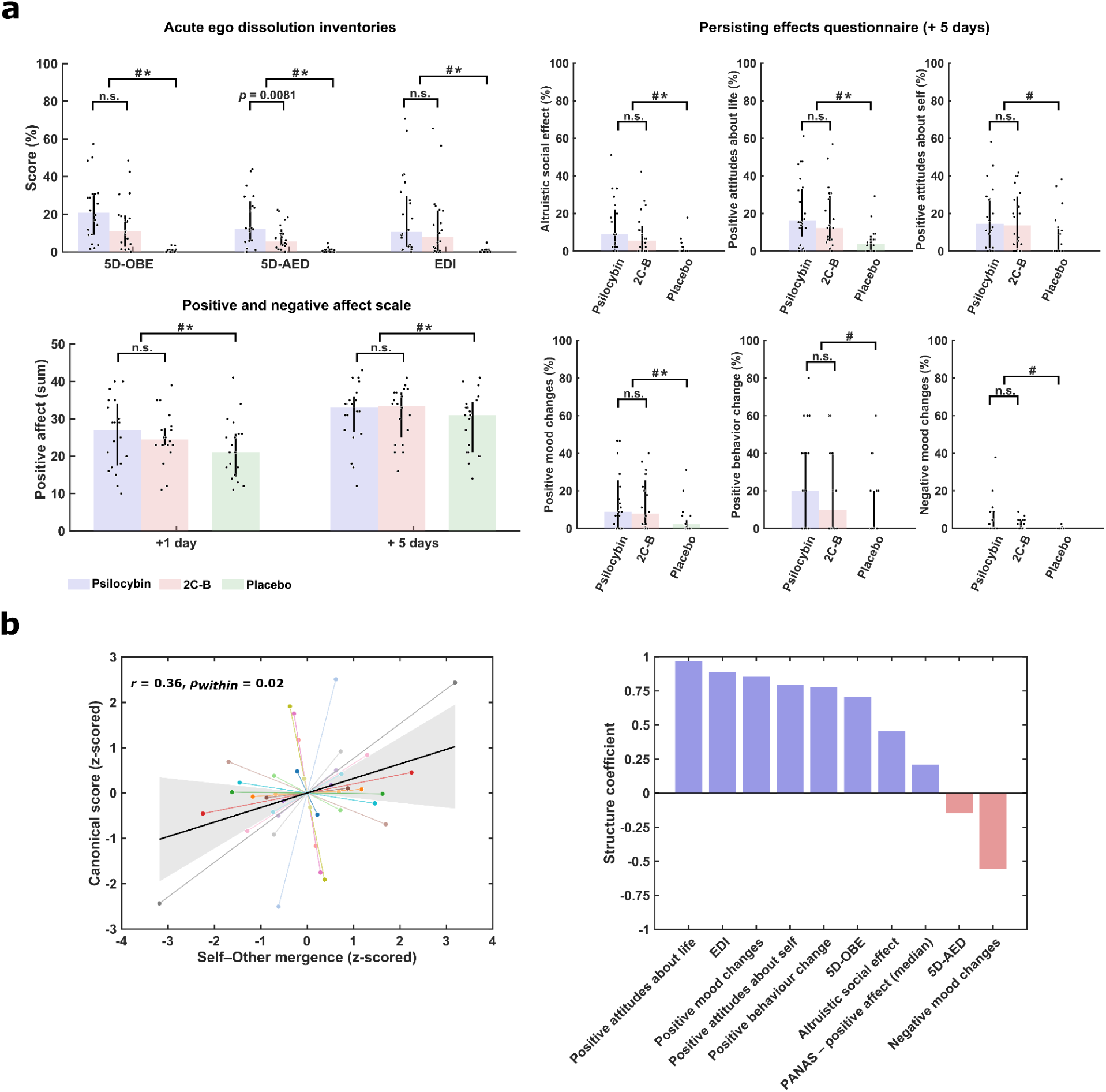
(Sub)acute psychosocial effects and their multivariate association with Self-Other mergence. **a**, Subjective effect outcomes. Acute ego dissolution was assessed using the retrospective 5-Dimensions of Altered States of Consciousness questionnaire (5D-ASC; oceanic boundlessness [OBE], anxious ego dissolution [AED]) and the Ego Dissolution Inventory (EDI). Positive and Negative Affect Scale (PANAS) scores were collected at + 1 and +5 days post-session, and persisting effects were measured using the Persiting Effects Questionnaire (PEQ) subscales at +5 days. Both psychedelics were associated with higher scores than placebo on several outcomes (see brackets), with significance assessed using Tukey-corrected pairwise comparisons. Bar graphs show the median +/- IQR; each dot represents one participant (N = 20). Significance (*p* < 0.05) per contrast is represented as the following; # = psilocybin vs placebo, * = 2C-B vs placebo, n.s. = not significant. **b**, Left: Multivariate association between Self-Other mergence and subjective outcomes. The scatter plot shows the repeated-measures association between Self-Other mergence and the canonical (sub)acute psychosocial response profile. Points are within-subject, drug-aware residuals (z-scored); segments connect distinct sessions from the same participant. The black line is the shared repeated-measures correlation fit and the shaded band shows its 95% CI (subject-level bootstrap). Because residualisation includes an intercept and drug term, placebo observations plot near the origin. Right: The bar chart depicts structure coefficients (correlation between each outcome and the canonical variate), indicating the relative contribution of each measure to the multivariate association.

Acutely, all ego dissolution dimensions were significantly elevated under both 2C-B and psilocybin, with the strongest effect in 5D-ASC oceanic boundlessness (F_1,39_ = 33.08, *p* < .0001). Psilocybin additionally produced greater elevations in dysphoric aspects of ego dissolution (5D-ASC anxious ego dissolution) relative to 2C-B (*p =* .0134, *d* = 0.94). Subacutely, a robust main effect of drug was observed across all positive valence dimensions of the PEQ, with altruistic prosocial effects showing the clearest enhancement at +5 days (F_1,39_ = 18.37, *p* < .0001) with no difference between compounds (*p* = .168). No effects emerged for antisocial negative effects (*p* = .691). Within negative valence measures, only negative mood showed a significant drug effect (F_1,39_ = 4.44, *p =* .0415), driven by elevated scores under psilocybin relative to placebo (*p* = .0215, d = 0.88), but not 2C-B (*p* = .163). PANAS positive affect paralleled the PEQ results, with a main effect of drug (F_2,95_ = 4.56, *p* = .0128) but no interaction with follow-up time (*p* = .713), nor significant differences between drugs (*p* = .939). By contrast, PANAS negative affect, IOS, and SCS-R showed no significant drug or drug × time effects (minimum *p* = .26). These results indicate that psychedelics consistently enhance psychosocial wellbeing subacutely, with psilocybin potentially producing greater dysphoric ego dissolution and transient increases in negative mood.

We examined whether individual differences in psychedelic-induced Self-Other mergence were associated with a coherent (sub)acute psychological response profile. To test this, we conducted a drug-aware permutation MANOVA (10,000 permutations), residualising average drug effects so that only between-participant variance was assessed. We included all subjective outcomes that showed a significant drug effect. A significant overall association between Self-Other mergence and the outcome set (Pillai’s trace = 0.64, p_perm_ = 0.0012) was identified. Canonical analysis (Fig. 3b) showed that Self-Other mergence correlated with the first canonical variate (r_in-sample_ = 0.36, p_perm_ = 0.0041). Importantly, a repeated-measures correlation confirmed that within-participant fluctuations in Self-Other mergence tracked corresponding changes in the canonical variate (p_within_ = 0.02). The effect also generalised under cross-validation (r_out-of-sample_ = 0.32, p_perm_ = 0.044; Fig. 3b), indicating that the canonical relationship was stable and predictive. Canonical weights and auxiliary descriptive univariate regression results (none surviving FDR correction) are reported in the Supplementary Materials. Structure coefficients indicated strong positive correlations for positive-valence and ego dissolution measures, and a strong negative correlation for negative mood and ego dissolution changes. Altogether, these findings suggest individuals with greater increases in Self-Other mergence under psychedelics also reported more positive psychosocial responses, independent of general drug effects.

### Psychedelic-induced Self-Other mergence does not reflect (sub)acute cognitive impairment

A central concern for interpreting any cognitive outcome in neuropsychopharmacology is task “impurity”. Could the observed effect on Self-Other mergence be more parsimoniously explained by general cognitive interference or drug-induced sub(acute) executive dysfunction? To address this, we leveraged task measures acquired in the same timeframe as the p-FBT (see Figure 4, Methods for additional information).

**Figure 4.**
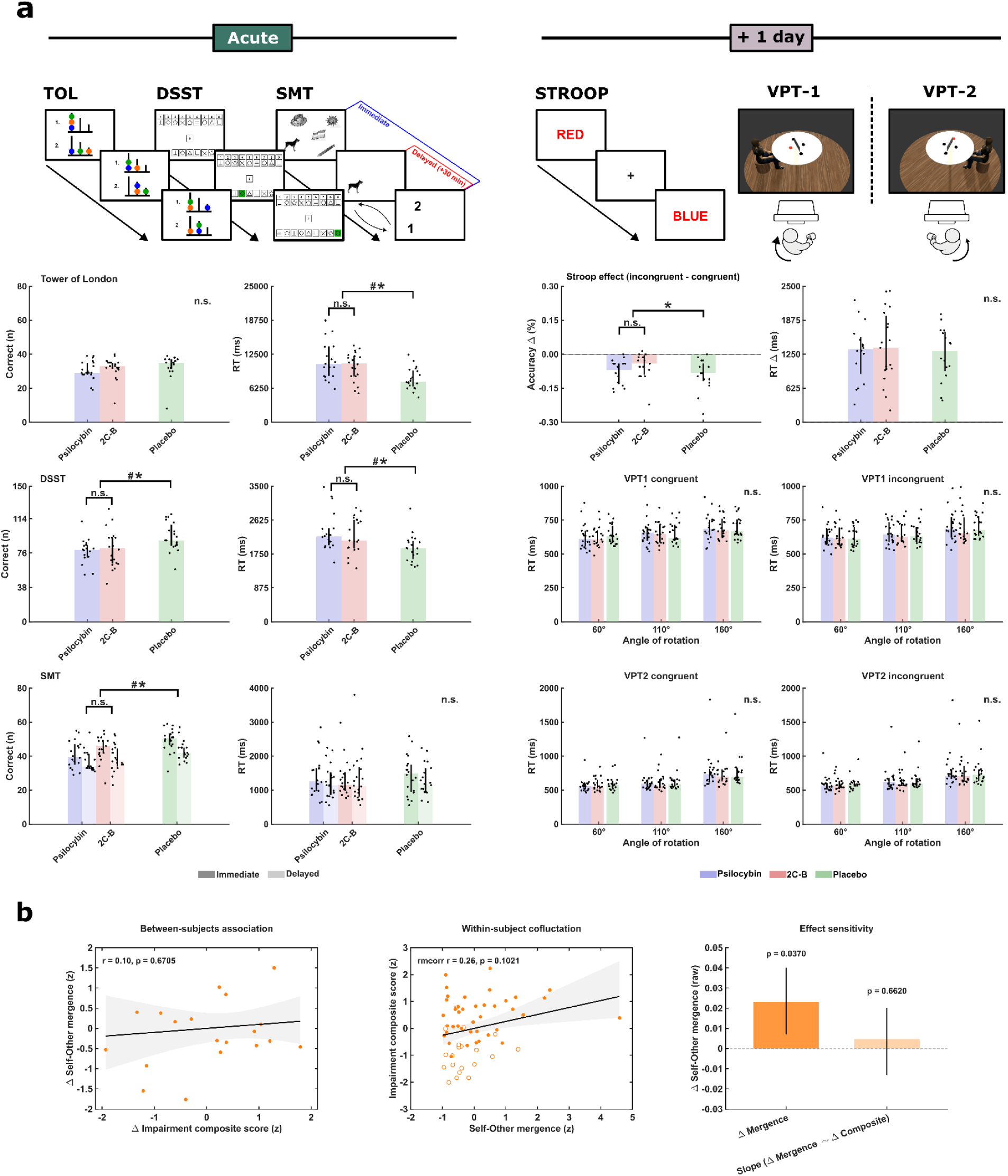
Assessing the confounding role of drug impairment. a, Task battery outcomes and approximate timings. The top panel of each column shows a schematic of each task as presented to participants, with its administration time relative to dosing. (1) Tower of London (TOL): participants judged whether the end arrangement (1 > 2) could be achieved in 2 to 5 steps, as quickly as possible. (2) Digit Symbol Substitution Test (DSST): participants matched novel inputs to the corresponding digit symbol pair within 90 seconds. (3) Spatial Memory Test: participants memorized the locations of a sequence of targets and indicated each target’s correct position (1/2); they were queried 30 minutes later about target locations. (4) Stroop task (STROOP): participants reported the ink colour of words relative to the congruent or incongruent named colour;. 6) Visuospatial Perspective taking Task (VPT): participants judged if a target was visible to the avatar (VPT-1, perspective-tracking) or whether a target was on the avatar’s left or right (VPT-2, perspective-taking); the avatar appeared at an angular disparity of 60°, 110°, or 160° relative to the participant, and body posture was manipulated to be congruent or incongruent with the direction of mental self rotation required on each trial. Bar graphs show the median ± IQR; each dot represents one participant. Significance (p < 0.05) per contrast is denoted as: # psilocybin vs placebo, * 2C-B vs placebo, n.s. not significant. All outcomes are presented and described in Tables S1. b, Relationships between impairment and Self-Other mergence. Left: between-subject association between change in impairment composite and change in mergence. Middle: within-subject repeated measures association between session-wise impairment composite and mergence across placebo and drug sessions (points show sessions per participant, with clear dots indicating placebo). Right: summary of the Δmergence mean and the sensitivity of Δmergence to Δimpairment (slope). P values are permutation based (10,000); shaded bands and error bars denote 95% CI (bootstrap).

On each acute dosing day, a cognitive task battery was administered. During peak drug effects, participants expectedly exhibited general executive impairment. On the Tower of London, response times were slower (F_1,38_ = 24.19, *p* < .0001) without a corresponding reduction in accuracy (F_1,38_ = 1.53, *p* = .2240). Response times were slower under both drugs relative to placebo (psilocybin vs placebo, *p* < .0001, *d* = 1.78; 2C-B vs placebo, *p* = .0040, *d* = 1.09), with no difference between drugs (psilocybin vs 2C-B, *p* > .0937). Digit symbol substitution test (DSST) performance showed marked reductions in accuracy, with fewer correct symbols (F_1,36_ = 29.83, *p* < .0001) under both drugs (psilocybin vs placebo, *p* < .0001, *d* = −1.83; 2C-B vs placebo, *p* = .0004, *d* = −1.36) and no difference between drugs (psilocybin vs 2C-B, *p* = .3424). DSST response times were also slower (F_1,36_ = 25.59, p < .0001) under both compounds relative to placebo (psilocybin vs placebo, *p* < .0001, d = 1.74; 2C-B vs placebo, *p* = .0014, *d* = 1.24), with no difference between drugs (psilocybin vs 2C-B, *p* = .2845). Spatial memory accuracy was reduced under both psychedelics for immediate and delayed outcomes (immediate: F_1,37_ = 17.41, *p* = .0002; delayed: F_1,36_ = 14.63, *p* = .0005). For the immediate phase, participants produced fewer correct items under psilocybin and 2C-B relative to placebo (psilocybin vs placebo, *p* < .0001, *d* = −1.63; 2C-B vs placebo*, p* = .0229, *d* = −0.88), with no definitive differences between the two (psilocybin vs 2C-B, *p =* .0710). The delayed phase showed the same pattern (psilocybin vs placebo, *p* = .0011, *d* = −1.31; 2C-B vs placebo, *p* = .0205, d = −0.93), with no difference between drugs (psilocybin vs 2C-B, *p* = .4674). Response times for both spatial memory phases were unchanged (immediate, *p* = .595; delayed: *p* = .792).

One day after dosing, we examined whether the observed shift in Self-Other mergence could reflect a residual impairment in the capacity to maintain distinct competing representations, rather than a specific change to Self-Other belief integration. We collected two control tasks administered in the same session as the pFBT probing this capacity at two levels: response conflict (Stroop), requiring participants to resolve stimulus conflict, and egocentric conflict (VPT), requiring participants to adopt another agent’s physical viewpoint under increasing spatial demand. Both being lower-level than the pFBT in their inferential demands, any non-specific drug effect on representational separation would be expected to manifest in both. On the Stroop task participants instead exhibited better interference control, demonstrated by reduced accuracy interference effects under drug post-administration (F_1,30_ = 6.39, *p* = 0.0169). This effect was driven by 2C-B (psilocybin vs placebo, p = 0.4428, ; 2C-B vs placebo, p = 0.0096, d = 1.11; psilocybin vs 2C-B, *p* = 0.1620). However, it should be noted that general accuracy scores were preserved and near ceiling (F_1,30_ = 0.33, *p* = 0.5695), and no changes were identified in response time interference effects (F_1,30_ = 0.33, *p* = 0.5695). Performance on the VPT showed clear sensitivity to task demands. Under placebo, response times increased with angular disparity, (F_2, 198_ = 107.04, *p* < .0001), and this disparity effect differed across blocks (task × disparity: F_2, 198_ = 18.03, *p* < .0001). In contrast, psychedelic administration did not increase interference. There was no evidence that drug status modulated congruency (drug × congruency: F_1, 663_ = 0.25, *p* = .6170; drug × task × congruency: F_1, 663_= 0.01, *p* = .9230). Analyses of the congruency cost score (incongruent minus congruent) were consistent with this pattern: there was no main effect of drug on RT cost, F_1, 319_ = 1.31, *p* = .2540, or accuracy cost, F_1, 319_ = 0.71, *p* = .4010, and drug effects within each block were not significant (RT cost VPT1 *p* = .2540, VPT2 *p* = .1360; accuracy cost VPT1 *p* = .4010, VPT2 *p* = .1410).

Using these findings, we examined three possible cognitive accounts of the observed effect on Self-Other mergence by computing a composite “impairment burden” score across all significant acute intensity (e_max_) and task outcomes. First, Δmergence did not track Δcomposite across participants (*r* = 0.10, p_perm_ = 0.6705). Second, within-subject co-fluctuation of impairment burden scores across sessions was not supported (*r* = 0.26, p_within_ = 0.1061). Third, the observed effect on Self-Other mergence remained statistically significant even after adjusting for Δcomposite (intercept = 0.023, p_perm_ = 0.037; slope p_perm_ = 0.6620). Independent correlations reported in the Supplementary Materials yielded the same overall pattern (all covariation tests non-significant). Taken together, these results that the drug-induced shift towards increased Self-Other mergence cannot be readily explained by sub(acute) changes to a participant’s broader cognitive abilities.

### Acute effective connectivity changes underlie persisting Self-Other mergence

Psychedelics disrupt the functional organisation of association cortices involved in self-referential processing, to produce a transient loss of coherent self-awareness^58^. To examine how such disruptions might underlie persisting Self-Other mergence, we lastly applied spectral dynamic causal modelling (DCM) to peak-effect 7T rsfMRI data. Focusing on an a priori Theory of Mind (ToM) network (Fig. 5a, Methods), we tested whether psychedelics alter the organisation of effective connectivity (EC) between core ToM regions in a manner consistent with altered belief updating.

**Figure 5.**
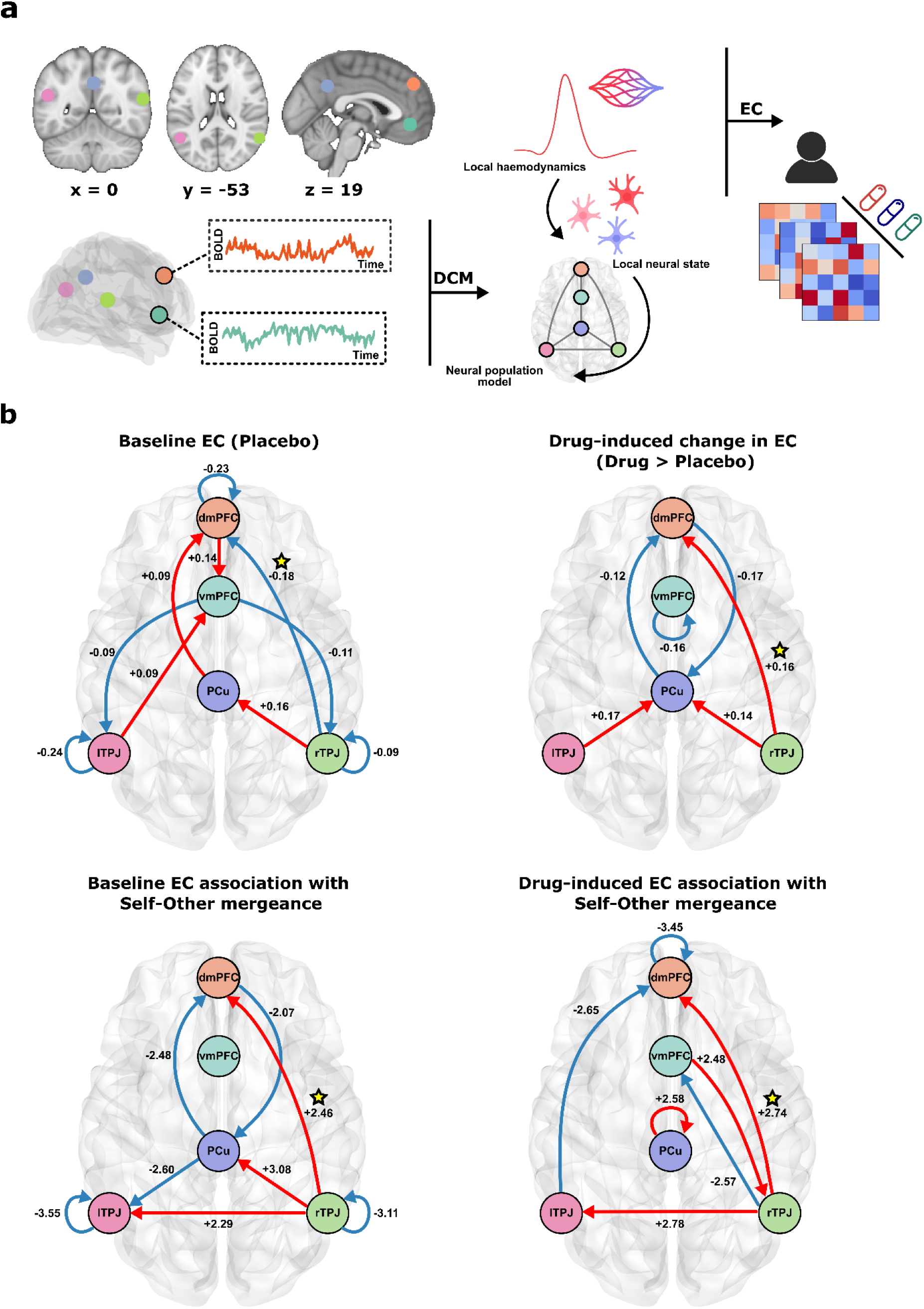
Dynamic causal modelling of effective connectivity (EC) under psychedelics and its association with Self-Other mergence. a, Theory of Mind network regions of interest (ROIs) are shown on canonical brain slices. Resting state fMRI data acquired approximately 110 min post administration were fitted using spectral DCM, which estimates directed effective connectivity (EC) between regions by modelling local haemodynamics and neural population dynamics. b, Four maps are shown, each visualising a different parameter from the hierarchical PEB model. ‘Baseline EC (placebo)’ shows posterior expectations of EC parameters in Hz, with red indicating positive (excitatory) and blue negative (inhibitory) influences. ‘Drug-induced change’ shows posterior expectations of the contrast [drug – placebo] in Hz, with red indicating an increase (increased excitation or decreased inhibition) and blue indicating a decrease (decreased excitation or increased inhibition) relative to placebo, regardless of the baseline sign. “Baseline EC association”shows regression coefficients linking baseline EC to the behavioural covariate (drug-induced change in Self Other mergence). “Drug induced EC association” shows regression coefficients linking the contrast EC change [drug –minus placebo] to the behavioural covariate. Self connections are log scaled parameters and are inherently inhibitory. Connections are displayed only when the posterior probability that the corresponding parameter is non-zero exceeds 0.99, applied separately for each map. Therefore, a connection can be absent in the baseline map yet present in the drug minus placebo map if there is stronger evidence for a change than for the baseline effect itself. ★ marks effects that converge across drug contrasts. Posterior probability and effect size matrices at a liberal threshold(>.5) are provided in the Supplementary Materials. Abbreviations: dmPFC, vmPFC, PCu, lTPJ, rTPJ.

Bayesian model reduction and averaging revealed very strong evidence (posterior probability > 0.99) for six connectivity parameters differing between drug and placebo conditions (Fig. 5b). At baseline (placebo), the network exhibited a mixture of excitatory and inhibitory influences. We will illustrate how to interpret these maps by taking one example connection, rTPJ→dmPFC . At baseline (placebo), group-level DCM estimates indicated an inhibitory connection from rTPJ to dmPFC. Under psychedelics, this connection exhibited an attenuation of inhibitory influence, consistent with relative disinhibition. Across participants, those with a more excitatory baseline connection (i.e. less inhibitory) exhibited larger drug-induced increases in Self-Other mergence (Λ). Furthermore, participants who showed the strongest excitatory shift in this connection under psychedelics also displayed the strongest behavioural evidence of increased Self-Other mergence 24 hours later. Exploratory analyses suggested that this attenuation was positively associated with the multivariate psychosocial response profile described above (see Supplementary Materials). Furthermore, specificity analyses using a TPJ-implicated task-active network disrupted by psychedelics, the salience/ventral-attentional network^59^, did not reproduce our results (see Supplementary Materials), suggesting functional specificity of this pathway. Together, these findings suggest that the rTPJ→dmPFC pathway is normally inhibitory, that psychedelics attenuate this inhibitory tone, and that the extent of this attenuation predicts the magnitude of behavioural change in Self-Other processing.

## Discussion

We show that 2C-B and psilocybin induce a state of Self-Other mergence. More specifically, there is an increase in sensitivity to agent-independent prediction errors, which encode a generic surprise signal, divorced from agent-identity i.e. (*me* or *you*). This shift, from Self-Other distinction to Self-Other mergence, persists beyond the acute drug phase, into the subacute *afterglow* period, and cannot be attributed to drug intensity nor residual changes to global cognitive abilities.

Human neuroimaging studies^8,10,11,60,61^ and electrophysiological studies in humans^9^ and non-human primates^62–66^ have found converging evidence that computational units of learning and belief formation can be encoded in anatomically segregated, agent-specific neuronal populations. This facilitates Self-Other distinction and provides a neurocomputational architecture for mentalising. It has previously been shown that this agent-specificity in prediction error encoding is malleable, and sensitive to social context^11^.

Our findings add to this literature by showing that pharmacological manipulation with psychedelics also reduces the agent-specificity of prediction errors. This is consistent with previous work showing that ketamine and psilocybin impair one’s ability to discriminate Self-generated from Other-generated touch^51^ and modulate bodily Self-experience^67^, and that psilocybin attenuates the distinctiveness of neural responses to hearing the voice of oneself versus another person^52^. Where these previous studies speak to Self-Other distinction with respect to observable physical phenomena (e.g. *Whose voice is that?* or *Whose touch is that?*), the current work extends this research to unobservable mental phenomena (e.g. *Whose belief is that?*).

A recent meta-analysis found that classical psychedelics enhance emotional empathy, the propensity to experience other people’s emotional states^50^. This finding has also been replicated in patients with major depression^43^. However, these previous studies found no effect of psychedelics on cognitive empathy, the ability to accurately infer and report the mental states of Others. Our findings show that the picture is more nuanced. Not only do psychedelics increase one’s propensity to attribute another person’s affective states to themselves, but also to attribute another person’s non-affective belief updates to themselves (*allocentric* belief updating). This is consistent with previous findings that psychedelics acutely make people more suggestible^68^ and more likely to adopt the opinions of Others^69^. In our data we found that behaviour was best explained by a model that included not only allocentric updating, but also *idiocentric* updating (propensity to attribute one’s own mental states to Other), and that these two vectors of belief updating were modified in equal measure. This drug-induced Self-Other mergence does not manifest as enhanced cognitive empathy, which would result in improved accuracy at identifying the beliefs of the other person. However, our findings indicate that blurring of Self-Other boundaries with respect to affective mental states extends to non-affective mental states. We therefore propose that the theory of psychedelics influencing affective empathy, and not cognitive empathy, is an oversimplification.

Our demonstration that 2C-B and psilocybin act on the processes underlying cognitive empathy has important implications for understanding the therapeutic effects of psychedelics in mental illness^70,71^. It is thought that the (sub)acute subjective effects of psychedelics, such as ego dissolution and perceived connectedness, are predictive of therapeutic efficacy^39,40^. Converging evidence from preclinical studies and human neuroimaging suggests that the enduring prosocial state during the subacute *afterglow* period may reflect a reopened critical period for social learning and cognitive restructuring^72–76^, necessary for the *integration* component of psychedelic-assisted psychotherapy^77,78^. This window of neuroplasticity appears to be induced by 5-HT_2A_-mediated increases in prefrontal excitatory signalling^79^. In our study we found that interindividual variability in psychedelic-induced Self-Other mergence, during this subacute period, was predictive of a canonical variate capturing both acute ego dissolution and subacute psychosocial wellbeing. This is consistent with previous work showing that people who exhibit less of a neural Self-Other distinction in reward processing, are more likely to engage in prosocial behaviour^80^.

We found that drug-induced changes to Self-Other mergence were positively associated with self-reported psychosocial wellbeing. The direction of this association is unlikely to be universal. It has previously been shown in healthy adults that a baseline propensity for Self-Other mergence is associated with transdiagnostic subclinical traits of mental illness^10^. Furthermore people with diagnosed borderline personality disorder (BPD) exhibit enhanced idiocentric updating, relative to the general population^27^. In healthy physiology, the brain is likely to balance Self-Other distinction with Self-Other mergence in a way that suits the current social circumstances^11^. We speculate that this ability to adaptively generalise between Self and Other, without over-relying on Self-Other mergence, is a marker of good mental health. The period of heightened plasticity following psychedelic administration may enable a re-learning of Self-Other boundaries, which could be facilitated by cognitive training or psychotherapy. Whilst, in our data, we observed heightened Self-Other mergence one day after drug administration, it will be useful for future work to establish trajectories of Self-Other boundary transformation over a longer period of time and to directly assess whether psychedelics are simply promoting Self-Other mergence, or promoting sensitivity to Self-Other boundary re-learning.

Psychedelics profoundly alter how people form beliefs about themselves and about the world, and this is not without risks^81^. In order to develop safe, individualised treatments in the emerging field of psychedelic psychiatry, it will be useful to have access to objective biomarkers that can predict and track treatment responses^82^. Quantifying Self-Other mergence in belief updating may provide a tool to predict how individuals will respond to psychedelic treatment. For instance, increased monitoring may be warranted during the subacute phase if giving psychedelics to individuals with maladaptive cognitions and behaviours related to an over-reliance on Self-Other generalisation, such as those exhibited by those with personality disorders, or individuals showing high Self-Other mergence in pre-treatment behavioural screening. Individual variability in Self-Other processing may also explain why some of the general population experience adverse effects or particularly positive psychosocial effects when using psychedelics recreationally.

Our findings also speak to a possible computational mechanism behind acute psychedelic-induced ego dissolution. In previous work applying reinforcement learning models to investigate the impact of 5-HT_2A_ agonists on learning and belief formation, it was shown that lysergic acid diethylamide (LSD) enhances learning rates in humans^83^ whilst 5-HT_2A_ antagonism decreases learning rates in rodents^84^. Recent work has also shown that LSD-induced learning rate enhancement is correlated with functional connectivity changes in the default-mode network (DMN) that are thought to reflect ego dissolution^85^, consistent with the REBUS (RElaxed Beliefs Under pSychedelics) model^86^. The REBUS model proposes that psychedelics reduce the precision of beliefs, leading to an upweighting of prediction errors and increased sensitivity to new information. In allowing the brain access to new hypotheses about reality, previously rigid beliefs underpinning one’s notion of Self may also be loosened, leading to ego dissolution^87^. Interestingly, the effect observed in the current study was not driven by enhanced sensitivity to surprise, but specifically enhanced sensitivity to cross-agent leaked surprise. By explicitly operationalising selfhood within our data and model, we found that psychedelics directly relax the constraints defining Self-Other boundaries in the belief-updating process. It is possible to conceptualise this within the REBUS framework by considering the neural instantiation of prediction errors as itself susceptible to a meta-learning process wherein metarepresentational learning signals^6^ are upweighted in the face of reduced precision of Self-Other boundaries. In any case, the dimension of altered belief-updating introduced in the current study may permit a direct and objective quantification of ego dissolution, beyond indirect physiological correlates.

Default-mode network (DMN) functional disintegration is a reported correlate of acute ego dissolution and enduring psychedelic effects^73,75,88^, and this is complemented by our neuroimaging findings. Using spectral dynamical causal modelling (DCM) to estimate effective connectivity between hubs of the so-called *social brain*, we found that 2C-B and psilocybin induced acute connectivity changes associated with subsequent Self-Other mergence. Most striking, was a drug-induced attenuation of an inhibitory connection from the right temporoparietal junction (rTPJ) to the dorsomedial prefrontal cortex (dmPFC), both components of the DMN. Both the rTPJ and dmPFC are involved in inferring the mental states of Others^89,90^. Previous work supports a role for the dmPFC in distinguishing Self from Other^91,92^; dmPFC neurons encode specific information about other agents’ behaviours^93^ and mental states^12^. At the same time, the rTPJ is thought to play a role in suppressing one’s current perspective, to allow for alternative perspectives to be considered^94–97^. Causal evidence from transcranial magnetic stimulation (TMS) and lesion mapping further corroborates this, with disruption of the rTPJ and dmPFC selectively impairing Self-Other boundary regulation in belief and decision-making^92,98–102^. It has similarly been recently shown that psilocybin-induced disembodiment is associated with altered rTPJ effective connectivity^103^. Taken together, the existing literature suggests that these two brain regions act in concert to allow for perspective-taking with selective attribution of a mental state to a specific agent. Our results show that when an inhibitory connection from rTPJ to dmPFC is weakened, mental states shift to become less agent-specific and more agent-independent. Moreover, individuals with weaker baseline rTPJ–dmPFC inhibition were more susceptible to psychedelic-induced Self-Other mergence, suggesting a potential biomarker for predicting treatment responsiveness. Together, these findings inform our understanding of how hubs of the *social brain* interact to modulate Self-Other distinction and how connectivity changes within this network relate to the subjective effects of psychedelics. These findings could be used to guide future work investigating adjunctive interventions (e.g., TMS) to probe causal mechanisms or modulate maladaptive belief updating^98,104^.

There are limitations to the present work. First, as the study was conducted in a small sample of healthy volunteers, we cannot determine whether the drug-induced changes we observed are clinically significant, nor can we exclude the possibility that effects would differ in patient or psychedelic-naive populations. Whilst our primary statistical contrast on Self-Other mergence was significant, individual compound effects were equivocal, with 2C-B showing comparable mean effects to psilocybin but greater within-subject response variability. Both 2C-B and psilocybin produced comparable effects on Self-Other processing and wellbeing at moderate doses, suggesting a generalisable mechanism across psychedelics, yet the absence of clear significant differences in this sample should not be taken as evidence that the two compounds act equivalently. Secondary pharmacodynamic distinctions between psychedelics have been shown to scale up to whole-brain functional connectivity ^105–107^, and may modulate the haemodynamic response function that underpins the core neurovascular coupling assumptions of biophysical models such as DCM^108^. Furthermore, our regions of interest were derived from resting-state data based on a meta-analytic identification of Theory of Mind circuitry, entailing a degree of reverse inference. Our control DCM analysis confirmed that generalised psychedelic-induced connectivity changes within salience circuitry could not reproduce the observed association with Self-Other mergence. However, the pFBT demands a range of cognitive faculties that clearly extend beyond just Theory of Mind computations. The interpretation of our findings should rely on the convergence across computational, behavioural, and effective connectivity outcomes, rather than on any one of these alone. Larger comparative studies and pharmacological challenge designs (e.g., selective 5-HT_2A_ antagonists, serotonin reuptake inhibitors) will be needed to specify the neuropharmacological basis of the effects reported here.

Finally, to interpret our results it is important to consider what it might mean for a computation to be social. Through one of the most productive debates currently running in social cognitive neuroscience, a growing body of work argues that regions of the social brain, and derived behavioural processes, are not contextually specialised for social stimuli, but computationally specialised for inference operations that become relevant across social and non-social contexts when demands align ^109^. A region can implement the same computation across both domains and still be preferentially recruited when social framing raises the precision of that inference. Mahmoodi and Rushworth extend this further^110^, arguing that computations underlying social interaction are exapted from non-social functions, such that the rTPJ and dmPFC track agent-directed inference regardless of explicit social framing, with social context modulating the magnitude of recruitment rather than engaging a categorically distinct processing mode. Under this view, the biological instantiation of Self-Other distinction may rely on neurocomputational architecture that did not evolve for social interaction specifically, but is selectively tuned by it. Our neurobehavioural findings, and prior validation of the effectiveness of the pFBT’s social framing and computational parameters^10,11^, perhaps speak to this level of description. A natural extension is to next ask how such computations scale to naturalistic dyadic exchange, where social context is not just a framing but a live generative process during which Self-Other boundaries are negotiated in real time.

In summary, our findings show that classical and novel psychedelics promote Self-Other mergence with respect to prediction errors, the fundamental computational units behind learning and belief formation. By integrating principles of computational psychiatry with psychedelic psychiatry, we have shown that there are objective, quantifiable brain-based and behaviour-based biomarkers of psychedelic-induced Self-Other mergence. These markers capture biological, psychological and social dimensions of an individual’s state. We propose they may be useful for holistically tracking the effects of a person’s psychedelic treatment. More broadly, our findings support the view that the neurocomputational basis of selfhood lies in flexible and manipulable Self-Other boundaries. We hope these insights encourage a shift away from treating selfhood as an ineffable quality and toward viewing it as a set of quantifiable, measurable processes.

## Methods

### Participants

The data were collected as part of a larger trial (trial register: NL8813, approved by the Academic Hospital and the University’s Medical Ethics Committee of Maastricht University (NL73539.068.20)^105,111^. Twenty-two healthy participants (11 female) aged 19-35 years (mean ± SD: 25 ± 4 years) were recruited by word of mouth and advertisement shared via Maastricht University social media. Participants were required to have had at least one previous lifetime psychedelic exposure (e.g., psilocybin, mescaline, LSD), but no exposure within the past 3 months. Those with psychiatric, major medical, endocrine, or neurological conditions, pregnant or lactating women, those not using reliable contraception, concomitant drug use, with a history of adverse reactions to psychedelics, with uncorrected or abnormal vision, or MRI contraindications were excluded (see Supplementary Materials for more details). All volunteers provided written informed consent before participation and were compensated for their participation time.

### Study design

We employed a double-blind, placebo-controlled, randomised crossover design with three acute rsfMRI sessions and two follow-ups per cycle (in-person at +1 day and remotely at +5 days; see Fig. 1). During dosing sessions, participants received 15 mg psilocybin, 20 mg 2C-B, or placebo (bittering agent). Doses were selected based on prior evidence of psychotropic equivalence, as detailed in companion manuscripts^111,112^. Acute eyes-open rsfMRI was acquired approximately 110 min after administration. In the present sample, real-time psychotropic equivalence was reasssesed immediately before and after rsfMRI acquisition across multiple measures, with no significant differences observed between compounds (see Supplementary Table 1). Further details are provided in the Supplementary Materials.

### The probabilistic false beliefs task (p-FBT)

One day after each dosing session (FU1) and following negative drug and alcohol screenings, participants returned to the laboratory to complete a probabilistic false belief task (p-FBT)^11,27^ in person (Fig. 1). In the p-FBT, participants observed a sequence of binary outcomes generated from a Bernoulli distribution with an underlying probability, *P*. The task was framed such that participants imagined themselves observing a sequence of sales in a tourist store on a tropical island, where each purchase was either a pink umbrella (with probability *P)* or a yellow sun-shade (with probability *1-P*). *P* could take any value between 0 and 1, and the task was designed such that *P* gradually drifted along a random walk. Through a series of successive *sampling trials,* participants were instructed to infer, and keep track of, the drifting hidden variable *P*.

In addition to this, participants were instructed to infer, and keep track of, another agent’s false beliefs about the drifting hidden variable *P*. The task included a fictional store manager who sampled only limited information, and sometimes misleading information, about the observed outcomes. Each sampling trial was *shared*, *privileged*, or *decoy*. In *shared sampling trials*, an icon of the cartoon manager looking through an open door indicated that the manager was also observing the outcome. In *privileged sampling trials*, an icon of the cartoon manager behind a closed door indicated that the manager was in a different room, unable to observe the information from the *sampling trial*. Finally, in *decoy sampling trials*, participants were told that the manager was observing sales on a security camera; however, unbeknown to the manager, the footage was out of date, and thereby uninformative about what the customers were really buying. These *decoy sampling trials* were indicated by an image of the cartoon manager’s face next to an icon of a computer disc.

Trial sequences were pre-generated such that the underlying probability trajectories for the participant (*P_Self_*) and the manager (*P_Other_*) were uncorrelated. The uncorrelated random walks were bound between 0 and 1 and adjusted in steps of 0.025 on each sampling trial. *Privileged sampling trials* drew outcomes from *P_Self_*, *decoy sampling trials* drew outcomes from *P_Other_*. In *shared sampling trials* the outcomes were randomly drawn from either probability distribution.

A total of 360 trial sequences were pre-generated, each containing 120 of each of the three *sampling trial* types in a randomised order. One of these trial sequences was randomly selected for each participant’s testing session. Each *sampling trial* was presented for 1250 ms, followed by a variable intertrial interval with a fixation cross on screen for 400–600 ms. Every four to nine *sampling trials*, participants were given one of two possible *probe trials*. In a *Self-probe* trial, participants were prompted to report their own belief about the probability that the next trial will reveal a pink umbrella (*P_Self_*). Alternatively, in *Other-probe* trials, participants were prompted to report an estimate of the manager’s false belief about this probability (*P_Other_*). These reports were made by positioning a cursor along a continuous probability scale. For *Other-probe* trials, participants were explicitly instructed to consider which sales the manager had seen or missed, including information viewed on the misleading security footage. Whilst not explicitly instructed to to maintain separate belief streams for Self and Other, participants were instructed to infer these two probabilities and keep track of them, as accurately as possible.

Prior to testing, participants received a detailed walkthrough of the task, including training runs and an introductory example. This process explained the different trial types and the use of the continuous rating scale. Participants who rated their understanding as *poor* or *very poor* were required to repeat the instructions and the practice phase. Participants were informed that the task was intentionally challenging and were encouraged to perform to the best of their ability. Each testing session began with a practice run of the task. Feedback was provided only during the introductory example; no feedback was given during the main task. To minimise fatigue, a self-paced break was introduced halfway through the task. The task was programmed using Matlab (MathWorks, Provo), visualised with Cogent 2000 (v125) and Cogent Graphics (v1.29), and administered in a quiet testing room.

### Quantifying task performance

For each behavioural testing session, we evaluated task performance by computing the Spearman correlation coefficient between the participant’s responses on *probe trials* and the true underlying Bernoulli parameters *P_Self_* and *P_Other_*. In addition, for each testing session we also computed a permutation-based null performance measurement, by randomly shuffling the subject’s responses on *probe trials* 1000 times, and each time computing the Spearman coefficient between the shuffled responses and the underlying Bernoulli parameters. We then subtracted the average of these null correlation coefficients from the true correlation coefficient. The resulting value represents behavioural performance, relative to chance-level random responding.

It is important to note that participants only sampled limited information about the true underlying probabilities *P_Self_* and *P_Other_*. It was therefore not possible for participants to make perfect inferences about *P*. We therefore approximated an “optimal” performance for each testing session by simulating a subject who inferred *P_Self_* and *P_Other_*by using a recency-weighted average of recently observed outcomes. We used a simplified version of the model described below (see ‘Behavioural model’) where the simulated subject used parallel belief updates for Self and Other, with a perfect memory and no behavioural stochasticity. Learning rate α was arbitrarily set to 0.15. Across all the testing sessions that were delivered to participants, these simulated “optimal” observers had a mean performance-relative-to-chance correlation of 0.57 (IQR: 0.51 - 0.63). We used the upper quartile of values to denote a window of “optimal” performance (0.63 - 0.75). Performance metrics should therefore be interpreted so that 0 represents completely random behaviour and scores in excess of 0.6 represent near-optimal performance.

Two participants were excluded from behavioural modelling as they consistently performed at or below chance-level in multiple sessions. This left 20 participants for inclusion in the modelling analyses. One participant only completed two sessions, with missing data for the placebo session.

### Behavioural model

We fitted each participant’s responses on *probe trials* with a simple reinforcement-learning style model, wherein the belief, *B* ∈ [0, 1], about *P* on any trial *t,* is updated proportionally to a *prediction error* (PE). On any given *sampling trial*, the PE is the difference between the observed outcome (1 or 0) and the previous belief state. To model agent-specificity, we used parallel learning processes for Self and Other:

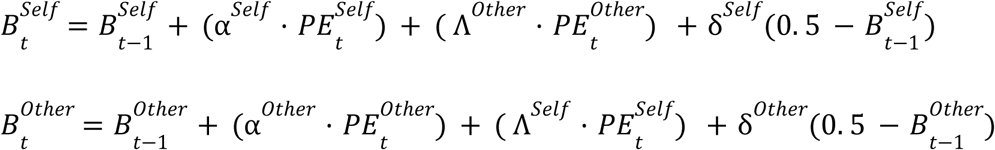

Here, *B*^*Self*^ denotes the participant’s belief, while *B*^*Other*^ denotes the participant’s simulation of the other agent’s belief. Thus, two belief representations are separately updated, based on observations available to Self and Other. α ∈ [0, 1] is a learning rate parameter, while δ ∈ [0, 1] is a memory decay rate, allowing beliefs to drift towards chance level. Importantly, updates for Self and Other may not be fully segregated and there may be a degree of Self-Other mergence. This is captured using *leak* parameter, Λ ∈ [− 1, 1], which allows one agent’s prediction error to erroneously inform the other agent’s belief. Where α describes sensitivity to surprise, Λ describes sensitivity to cross-agent leaked surprise. If Λ = 0 then prediction errors are entirely agent-specific. As Λ deviates from 0, prediction errors become progressively more agent-independent, resulting in Self-Other mergence of belief-updating channels. This form of model has previously been shown to fit behaviour well on the p-FBT ^10,11,27^.

In order to keep belief states naturally bound between 0 and 1, a factorised version of the model is fit to the data^27^. In this factorised form, Λ (sensitivity to cross-agent leaked surprise) is obtained by multiplying α with λ ∈ [− 1, 1].

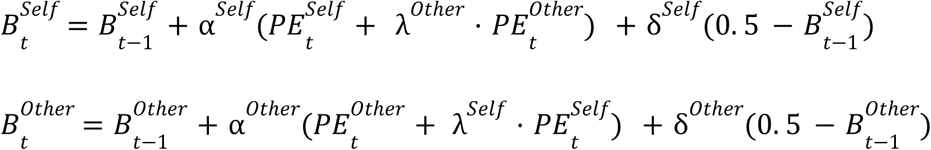

### Model fitting procedure

The model yields point estimates for *B*^*Self*^ and *B*^*Other*^ on each trial. These estimates were used to derive the likelihood of a participant’s responses across all *probe trials*. The likelihood was approximated using two Beta distributions (bound between 0 and 1), with the distribution mode set to the model-derived point estimate (*B*^*Self*^ or *B*^*Other*^) and the Beta distribution variance set to a participant-specific free parameter, τ^*Self*^ or τ^*Other*^, which can be thought of as decision temperature parameters, governing response stochasticity. High values in these parameters result in random responding, whilst lower values result in precise responses that are tightly coupled to the underlying belief states *B*^*Self*^ and *B*^*Other*^ .

The model can be specified in a variety of ways, resulting in a family of nested models that can be fitted to the data. Learning rate can be constrained such that α = α or they can be modelled as two separate free parameters. Temperature τ can be constrained such that τ*^Self^* = τ^*Other*^ or modelled as two separate free parameters. Memory decay δ can be constrained such that δ*^Self^* = δ^*Other*^ or modelled as two separate free parameters, or δ can be removed completely such that δ*^Self^* = δ^*Other*^ = 0. Finally, leak λ can be constrained such that λ*^Self^* = λ^*Other*^ or can be modelled as two separate free parameters, or removed completely such that λ*^Self^* = λ^*Other*^ = 0. In summary, has one or two parameters; τ has one or two parameters; δ has zero, one, or two parameters; λ has zero, one, or two parameters. This results in 36 nested model variants.

We fitted all 36 model variants to each individual testing session (three testing sessions per participant). Parameters were bounded through sigmoid transformation and fitted in inverse sigmoid space. Both α and δ parameters were bounded between 0 and 1, λ between -1 and 1, and τ between 0.001 and 0.08 (this upper bound on τ produces a near-uniform Beta distribution, corresponding to random responding). For each participant we sought model parameters with maximum posterior probability, given the data. We used fixed Gaussian priors over parameters and assumed a uniform prior over models. For each model variant, we calculated the Bayesian model evidence *P*(*x*|*M*) (probability of data *x*, given the model M), which favours models that provide a closer fit to the data, whilst penalising model complexity. Bayesian model evidence was calculated by marginalising the joint probability *P*(*x*, θ|*M*), with respect to the parameters θ. The exact calculation for *P*(*x*|*M*) is given by:

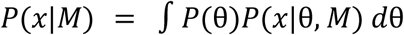

As per Story et al.^27^, we approximated this integral numerically, by randomly sampling parameters from the prior distribution, such that:

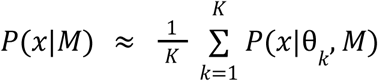

Where θ_*k*_ is a randomly sampled set of parameter values for a given model, and K is the total number of samples, set to 2000. Raw fitted parameter values in both unconstrained native space and transformed space are shown in supplementary figure S1a.

To measure parameter recovery for the best fitting model, we fitted the model to data generated for 1000 simulated participants. Simulated participants’ parameters were randomly sampled from the observed maximum a posteriori parameter estimates. Each parameter was sampled independently, with replacement. We computed a Pearson correlation coefficient between the generative parameters and fitted parameter estimates.

### Inference on parameters

To determine non-specific psychedelic-induced effects on parameters we specified the following mixed effects model:

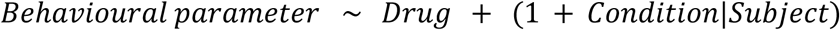

This model includes fixed effects for *Drug* (a binary variable which specifies whether the condition was a drug condition or a placebo condition), as well as random intercepts and random slopes for *Condition* (psilocybin, 2C-B or placebo) by *Subject*.

### Subjective effects

On each dosing day, psychotropic dose equivalency was reassessed using 0-100 visual analogue scales (VAS) to measure subjective high and effect intensity. The phenomenological content of each dosing visit was retrospectively evaluated using the 5 Dimensions of Altered States Questionnaire (5D-ASC) and Ego Dissolution Inventory (EDI) around 360 minutes post-drug intake^113,114^. Participants were contacted to assess “afterglow” effects at +1 day (in person) and +5 days (remotely via Qualtrics XM12). At both follow-ups, participants completed the Social Connectedness Scale–Revised (SCS-R) and the Inclusion of Other in the Self (IOS) to capture changes in social functioning^115,116^, as well as the Positive and Negative Affect Schedule (PANAS) to index ongoing mood states^117^. At + 5 days, they also completed the Persisting Effects Questionnaire (PEQ) to evaluate longer-term psychosocial changes attributable to treatment^118^. All outcomes were assessed by linear mixed effect models (LMEMs) using drug condition as a main fixed effect (and where applicable an interaction of time) and participant as a random intercept. Follow-up assessments were performed using Tukey’s method.

### Behavioural associations

To test whether drug-induced changes in Self-Other mergence covaried with (sub)acute psychosocial outcomes, we conducted a subject-level permutation MANOVA. Outcome measures were residualised for mean drug effects (active vs. placebo) and z-scored, ensuring that associations reflected within-person, drug-evoked covariation. Multivariate association was evaluated with Pillai’s trace, with significance determined from 10 000 within-subject permutations. Canonical correlation analysis then identified the linear outcome combination most strongly associated with Self-Other mergence, with significance tested by a 10 000-permutation Pearson correlation. A repeated-measures correlation further quantified the within-subject association between Self-Other mergence and canonical scores across conditions^119^. Robustness was evaluated using five-fold cross-validation, testing whether canonical scores could be predicted in held-out folds, with significance assessed against a 10 000-permutation null. This within-subject framework ensures that associations reflect how individual differences in Self-Other mergence track psychosocial responsinges under psychedelics, rather than being driven by overall drug–placebo differences.

### Cognitive interference testing

A standardised computer-based task battery (Eprime) was implemented during peak subjective effects to determine the impact of psilocybin and 2C-B across cognitive domains ^111^. The executive function arm comprised the Digit Symbol Substitution Test (DSST; processing speed and executive efficiency), Tower of London (TOL; planning and decision making), and the Spatial Memory Test (SMT, immediate and delayed +30 minutes; spatial working and episodic memory). Distinct task versions were used across sessions to minimise learning effects, and participants were refamiliarised with instructions prior to each task. Full task details and outcomes are defined in the Supplementary Materials. All outcomes were assessed by LMEMs using drug as a main fixed effect (and where applicable an interaction of time) and participant as a random intercept. Follow-up assessments were performed using Tukey’s method.

On the same day as the p-FBT (one day post-administration, FU1), we assessed potential residual impairment effects on cognitive interference and executive control using two additional computer-based tasks. These measures were included to evaluate whether drug-related effects on the pFBT could be explained by increased conflict sensitivity, reduced cognitive control, or broader residual impairment. Participants completed a standard 144 trial (50% congruency split) colour–word Stroop task (Neurobehavioral Systems, Presentation) implemented to quantify domain-general conflict interference and cognitive control ^120^. The primary outcomes were the congruency effect (incongruent-congruent) of mean response time (RT) and accuracy. Outcomes were assessed by LMEMs using drug as a main fixed effect and participant as a random intercept. Follow-up assessments were performed using Tukey’s method. The visuospatial perspective taking task (VPT, Unity engine) indexed interference during a social perspective conflict, with performance requiring resolving egocentric versus allocentric viewpoints (especially in Level 2), with additional interference manipulated by embodied congruency and angular disparity. ^121,122^ Participants viewed a table scene with a randomly generated avatar seated at one of six positions around the table (60°, 110°, or 160° left or right). On each trial, one of four discs around an occluder turned red as the target. In Level 1 (perspective tracking), participants judged whether the avatar could see the target (yes or no). In Level 2 (perspective taking), participants judged whether the target was on the avatar’s left or right. For the embodied manipulation, participants swivelled their chair to a marked spot so body orientation was congruent or incongruent with the required mental self rotation, maintaining gaze on the monitor and responding only once stationary. The task comprised 14 alternating mini blocks of 24 trials, preceded by 12 practice trials (6 per condition). Following ^100^, drug effects on performance were tested with trial-level LMEMs on participant level mean RT (correct trials) and accuracy, with fixed effects for drug, task (VPT1 vs VPT2), disparity (60°, 110°, 160°), and congruency, including the corresponding two-way and three-way interaction terms (for example drug × congruency × disparity).

### Confound control

To test whether psychedelic related increases in Self Other mergence could be better explained by domain general cognitive interference or acute drug intensity, we ran a confound sensitivity analysis. We built an “impairment burden” composite as the mean of direction aligned, z scored task and intensity measures, where higher values indicate worse performance or stronger acute effects. We computed deltas as drug minus placebo for both Self Other mergence and the impairment burden composite, then ran three tests: (i) between participant covariation between Δmergence and Δcomposite, (ii) whether the mean Δmergence persisted after adjustment, defined as the intercept from a regression of Δmergence on mean centred Δcomposite, and (iii) within participant session level co movement between mergence and the session composite using repeated measures correlation. Permutation based p values used sign flip inference throughout (10,000 permutations). Independent Δ correlations and composite contributions are reported in the Supplementary Materials.

### Administration order effects

To examine potential order effects, we estimated subject fixed-effects regressions for all significant task and questionnaire outcomes, including an interaction term between drug condition and session day (1–3). Cluster-robust standard errors were applied. No significant drug × day interactions were detected (all F_2,19_ < 1.7, p > 0.21).

### Neuroimaging acquisition

Eyes-open rsfMRI data were acquired using a 7T Siemens Magnetom scanner (Siemens Medical, Erlangen, Germany) equipped with a 32-channel Nova Medical head coil (Nova Medical Inc., Wilmington, MA). Functional images (516 volumes, approximately 12 minutes) were obtained using a gradient echo-planar imaging (EPI) sequence with 1.5 mm isotropic voxels (TR = 1400 ms). Preprocessing was performed using SPM12 employing a previously described effective connectivity pipeline^42,123^ adapted for ultra-high-field imaging. Steps included non-steady-state volume discarding, susceptibility distortion correction, slice-timing correction, realignment, spatial normalisation to the Montreal Neurological Institute (MNI) EPI template, and spatial smoothing with a 4-mm full width at half maximum Gaussian kernel. No significant differences in mean framewise displacement (FD, < 0.5 mm) were identified between conditions. Additional details are provided in the Supplementary Materials.

### Extraction of region coordinates

To evaluate whether dynamical changes in Theory of Mind (ToM) brain systems reflected acute psychedelic effects and subsequent alterations in mentalising abilities, we defined five regions reported as central to self-other processing such as belief attribution, perspective-taking, and understanding social intentions^124–126^. For this purpose, we isolated the dorsomedial prefrontal cortex, ventromedial prefrontal cortex, precuneus and the bilateral temporal parietal junction as targets (dmPFC, vmPFC, Pcun, lTPJ, rTPJ respectively). Neurosynth, a large-scale automated synthesis of functional neuroimaging data, was used to identify corresponding peak activation coordinates^127^, from which ROIs were centred and masked by an 8-mm radius sphere (see Table 1). More details can be found in the Supplementary Materials.

**Table 1.** Coordinates of regions of Interest. Abbreviations: dorsomedial prefrontal cortex (dmPFC), ventromedial prefrontal cortex (vmPFC), precuneus (Prcun), left temporoparietal junction (lTPJ), right temporoparietal junction (rTPJ).

| Region | MNI coordinates |  |  |
| --- | --- | --- | --- |
|  | X | Y | Z |
| Dorsomedial Prefrontal Cortex (dmPFC) | -2 | 42 | -8 |
| Ventromedial Prefrontal Cortex (vmPFC) | 2 | 46 | 38 |
| Precuneus (PCu) | 2 | -52 | 36 |
| Left Temporoparietal Junction (ITPJ) | -50 | -56 | 22 |
| Right Temporoparietal Junction (rTPJ) | 58 | -56 | 18 |

A general linear model was used to regress head motion and physiological noise from each regional time series using I) 6 head motion parameters (3 translation and 3 rotational), II) cerebrospinal fluid (CSF) signals (extracted from left ventricle using a 4-mm sphere [0 -40 -5]), and III) white matter signals (extracted from pons using a 4-mm sphere [0 -30 -25]). Low-frequency signal drifts were filtered using a 128-s high-pass filter. Global signal regression was not performed as there is evidence that it does not substantially impact results in small network analyses^128^.

### Effective connectivity estimation using spectral DCM

We fitted a spectral dynamic causal model (DCM) to each rsfMRI timeseries, using the DCM toolbox in SPM12. Spectral DCM fits a biophysical state-space model to the observed cross-spectra of BOLD signals, to estimate underlying neuronal states and the rate of change in neural activity in each region (in Hertz) as a function of activity in other regions^129,130^. For each of the 20 participants with p-FBT data, and each experimental session, a fully connected DCM was constructed using the five ROIs outlined in Table 1, with no inclusion of exogenous inputs. The DCM fitted the data well, and the amount of explained variance, averaged 88.13% (min = 79.08%, max = 96.26%) nor significantly differed across conditions (See Supplementary Materials). In DCM, self-connections are log-scaled and inherently inhibitory, reflecting a region’s sensitivity to incoming inputs: positive values indicate increased self-inhibition, while negative values reflect disinhibition and enhanced synaptic gain. Note that only self-connections are log-transformed (see Supplementary for details).

### Group inference using Parametric Empirical Bayes (PEB)

For each participant, three first-level DCMs (one per session) were summarised with a subject-specific second-level PEB model^131^. This 2nd-level model included an intercept and a drug regressor (2C-B and psilocybin vs. placebo). Group-level inference was then performed using a PEB-of-PEBs approach^132^, where subject-specific PEBs served as inputs to a third-level model. The third-level design matrix contained two regressors: (i) a constant modelling group-average connectivity, and (ii) a behavioural covariate reflecting drug-induced changes in Self-Other mergence (mean psychedelic minus placebo). This framework therefore allowed us to estimate group-average placebo connectivity, group-average drug effects, and associations between connectivity (both baseline and drug-induced changes) and behaviour. Models were pruned using exploratory Bayesian model reduction and Bayesian model comparison, employing an automatic greedy search over reduced models that iteratively removed parameters not contributing to model evidence. A Bayesian model average was then computed across the 256 best-fitting reduced models (default) from the final iteration. We applied a conservative statistical threshold of a posterior probability >0.99 equivalent to a Bayes Factor of >150 and considered a “very strong” evidence^133^. In the case of behaviour, the posterior probabilities express the likeliness of association between effective connectivity estimates and behavioural scores. The details of the biophysical model in DCM, model inversion at first-level and higher levels, and Bayesian model reduction, have already been extensively documented^132,134,135^. Further specifications are provided in the Supplementary Materials.

## Data availability

Code generated for analysis is to be made available at the following link: (https://github.com/PabloMallaroni/project_efc_afterglow). Data and accompanying covariates can be made available to qualified research institutions upon request to J.G.R and a data use agreement executed with Maastricht University.

## Author contributions

P.M. contributed to conceptualisation, investigation, methodology, formal analysis, data curation, project administration, visualisation, and writing (original draft, review and editing); N.L.M. contributed to supervision, investigation, project administration and writing (review and editing); K.H.P. contributed methodology and writing (review and editing); A.R. contributed methodology, and writing (review and editing); S.E. contributed to supervision, conceptualisation, methodology, formal analysis, visualisation and writing (original draft, review and editing); J.G.R. contributed to supervision, funding acquisition, project administration, data curation and writing (review and editing).

## Acknowledgements

We thank Emma de Brabander, Vitea Incerti-Medici, Ajna Jansson and Chantal Delaquis for their assistance with data collection, and Richard Benning and Erik Bongaerts for adapting the VPT for the Unity Engine. We are grateful to Dr. Cees van Leeuwen and Dr. Riccardo Paci for conducting medical screenings.

