## Supplementary Materials for "Neurocomputational evidence of sustained Self-Other mergence after psychedelics"

### Supplementary Materials for:
### **Neurocomputational evidence of sustained Self-Other mergence after psychedelics**

Pablo Mallaroni[1,2], Natasha L. Mason[1], Katrin H. Preller[3], Adeel Razi[4,5,6], Sam Ereira[7*] & Johannes G. Ramaekers[1*]

*These authors contributed equally to this work

[1]Department of Neuropsychology and Psychopharmacology, Faculty of Psychology and Neuroscience, Maastricht, the Netherlands
[2]Department of Computing, Faculty of Engineering, Imperial College London, London, United Kingdom
[3]Department of Adult Psychiatry and Psychotherapy, Psychiatric University Clinic Zurich, University of Zurich, Zurich, Switzerland
[4]Turner Institute for Brain and Mental Health, School of Psychological Sciences, Monash University, Clayton, Victoria, Australia
[5]Queen Square Institute of Neurology, University College London, London, United Kingdom
[6]CIFAR Azrieli Global Scholars Program, CIFAR, Toronto, Ontario, Canada
[7]Wolfson Institute of Population Health, Queen Mary University of London, London, United Kingdom

**Corresponding author emails***


**Table of contents:**


- **S1. Methods: Participants**
- **S2. Methods: Testing procedures**
- **S3. Methods: Drug administration**
- **S4. Methods: Self-report questionnaires and cognitive tasks**
- **S5. Methods: MRI acquisition**
- **S6. Methods: Meta-analytic region-of-interest coordinate extraction**
- **S7. Methods: Dynamic causal modelling (DCM)**
- **S8. Results: pFBT parameter retrieval *(Figure S1)***
- **S9. Results: Behavioural associations *(Figure S2, S3)***
- **S10. Results: DCM quality control *(Figure S4)***
- **S11. Results: Effective connectivity matrices *(Figure S5)***
- **S12. Results: Exploratory DCM multivariate associations *(Figure S6)***
-
- **S13. Results: DCM specificity analyses *(Figure S7)***
- **S14. References**

**S1. Methods: Participants**

Eligible participants were required to be between 18 and 40 years old, have prior experience with a psychedelic substance but not within the last three months, and have a body mass index (BMI) between 18 and 28 kg/m². Additional requirements included being medication-free (defined as no prescribed drugs for medical conditions), in good overall physical health, and free from major medical, neurological, or endocrine disorders. All participants were required to provide written informed consent. Individuals with prior psychedelic experience were specifically recruited to reduce the risk of severe adverse psychological reactions during ultra-high field 7T MRI acquisition.

Exclusion criteria comprised a personal history of psychiatric disorders, drug abuse or addiction, pregnancy or breastfeeding, lack of reliable contraception, previous serious adverse reactions to psychedelics, and standard contraindications for MRI. Before enrolment, participants underwent a screening by an independent study physician. This assessment included a medical history and physical examination, resting electrocardiogram (ECG), and laboratory testing (blood and urine samples analysed for haematology, clinical chemistry, and urinalysis). All participants received detailed information about the study procedures, potential side effects, rights and responsibilities, and their freedom to withdraw at any time without consequences. Written informed consent was obtained before inclusion, and participants received financial compensation for their time.

Initial eligibility was assessed via an online questionnaire (Qualtrics XM12), which captured demographic information, prior substance use, and geographic location. Those meeting preliminary criteria were invited to a remote briefing session outlining study procedures and assessments. After providing consent, participants completed structured inventories assessing psychiatric history (based on DSM Axis I and WHO ICD-10 classifications) and drug use patterns. Candidates who remained eligible were scheduled for an in-person medical screening. During this session, the study psychiatrist verified all reported information through a detailed clinical interview, assessed physical and mental health, measured vital signs and weight, and recorded an ECG. A comprehensive physical examination was also performed, along with laboratory testing including urine toxicology for illicit substances, urinalysis, serum chemistry, haematology, and liver function measures. Participants were explicitly informed that although they might not personally benefit from the study, their participation could help generate knowledge with potential benefit for others. All volunteers were compensated for their time and participation; remuneration was not contingent on task performance.

**S2.Methods: Testing procedures**

Participants were instructed to abstain from psychoactive substances throughout the study. Specifically, they were required to refrain from psychedelic drug use for at least three months prior to each session, alcohol for a minimum of 24 hours, and other drugs of abuse for at least seven days before testing. On dosing days, participants were also asked to avoid caffeine and nicotine. Compliance was verified at each in-person visit with urine toxicology screening and a breathalyser test, and female participants additionally completed a pregnancy test. Use of any psychoactive substances outside of those administered as part of the study protocol was prohibited during the seven-week study period. Each dosing session lasted approximately seven hours. Participants were instructed to eat a light breakfast before arrival. If all screening results were negative, dosing commenced. During the experimental session, participants remained under continuous supervision and were permitted to rest, read, listen to music through headphones, or converse with study staff. At the end of the day, investigators confirmed that participants were fit to leave. Follow-up procedures included an in-person visit the next day and an online assessment five days later, during which participants completed self-report inventories of persisting psychedelic effects (via Qualtrics (XM12). Participants were informed that this was a mechanistic study rather than a therapeutic study. Accordingly, no preparatory sessions with therapeutic framing or formal post-session integration were provided. Instead, preparatory discussions focused on ensuring understanding of study procedures and safety protocols, while research staff maintained close contact and remained available for direct support throughout all testing cycles.

**S3. Methods: Drug administration**

In the absence of existing 5-HT$_{2A}$ receptor occupancy data for 2C-B and a complex polypharmacology, moderate "equivalent" doses of 20 mg 2C-B and 15 mg psilocybin were approximated according to their psychotropic equivalence as defined by subjective drug high and intensity, extrapolated from prior study data. For the full rationale and evidence of this equivalence, see the companion manuscripts (Mallaroni et al., 2023; Mallaroni, Singleton, et al., 2024). Synthetic (powder) formulations of 2C-B and psilocybin were employed. 2C-B was obtained from Duchefa Farma B.V., Haarlem, Netherlands. Psilocybin was obtained from THC Pharm GmbH, Frankfurt, Germany. A permit for obtaining, storing, and administering 2C-B and psilocybin was obtained from the Dutch Drug Enforcement Administration. Participants were informed before the study that they would receive 20 mg 2C-B, 15 mg psilocybin, or a placebo on three separate occasions. Each intervention was administered orally, in a closed cup, either containing 200 mL of the vehicle bitter lemon (a bittering agent, placebo) or bitter lemon and psilocybin/2C-B (powder) to mask any potential taste. Blinding was assessed retrospectively at the end of each dosing day for which outcomes are reported in the companion manuscript (Mallaroni et al., 2023).

**S4. Methods: Self-report questionnaires and cognitive tasks**

Subjective effects were assessed repeatedly using visual analogue scales (VASs) at baseline (0h), +0.5, +1, +1.5, +2, +3, +4, +5, and +6 hours after drug administration (Holze et al., 2020). These scales included the primary VAS items: “any drug effect, and “drug high”, presented as 0-100 horizontal lines (%), marked from “not at all” on the left to “extremely” on the right . The VAS “any drug effect” is an overall effect measure to characterise the overall effect intensity and time course. Prior dose-effect studies have previously demonstrated it to be a useful marker for pharmacokinetic-pharmacodynamic modelling of psilocybin’s effects (Holze et al., 2023). Separately, “any drug effect” is interrelated with “drug high”, a measure of stimulating effects. These items were used to define dosage equivalence from previous data (Mason et al., 2020; Papaseit et al., 2018) and found to yield psychotropic equivalence in the complete behavioural sample across a range of measurements (Doss et al., 2024; Mallaroni et al., 2023; Mallaroni, Singleton, et al., 2024).

The 5D-ASC (Studerus et al., 2010) consists of 94 retrospective 0-100 VAS. The ASC items are grouped into five main dimensions comprising 11 lower-order scales (1) ‘oceanic boundlessness’ (OB) measures derealisation and depersonalisation accompanied by changes in affect ranging from heightened mood to euphoria and/or exaltation as well as alterations in the sense of time. The corresponding subfactors include “experience of unity,” “spiritual experience,” “blissful state,” “insightfulness,” and “disembodiment.” (2) ‘anxious ego dissolution’ (AED) measures ego disintegration associated with loss of self-control, thought disorder, arousal, and anxiety. The corresponding subfactors comprise “impaired control of cognition” and “anxiety.” (3) ‘visionary restructuralisation’ (VR) refers to ‘elementary hallucinations’, ‘visual (pseudo-) hallucinations’, ‘synaesthesia’, ‘changed meaning of percepts’, ‘facilitated recollection’, and ‘facilitated imagination’. It consists of the lower-order scales “complex imagery,” “elementary imagery,” “audio-visual synesthesia,” and “changed meaning of percepts.” (4) ‘auditory alterations’ (AA) refers to acoustic hallucinations and distortions in auditory experiences and (5) the dimension ‘reduction of vigilance’ (RV) relates to states of drowsiness, reduced alertness, and related impairment of cognitive function. The 5D-ASC is the most widely used psychometric assessment of classical psychedelic and entactogenic effects and has been deployed across a range of altered states of consciousness (Schmidt & Berkemeyer, 2018).

The Ego-Dissolution Inventory is a self-reported scale used to specifically assess subjective feelings of ego-dissolution/loss of self after drug intake (Nour et al., 2016). The questionnaire consists of 8 VAS items (0-100) which participants have to rate retrospectively, including: I experienced a dissolution of my “self” or ego; I felt at one with the universe; I felt a sense of union with others; I experienced a decrease in my sense of self-importance; I experienced a disintegration of my “self” or ego; I felt far less absorbed by my own issues and concerns; I lost all sense of ego; All notion of self and identity dissolved away.

The IOS, originally designed to measure the inclusion of romantic partners in self-construal, was adapted for this study. The 6-item scale presents seven images of two circles with varying degrees of overlap, representing "self" and "other” (Aron et al., 1992). Participants selected the image best describing their current relationship with others in general, with higher scores indicating greater connectedness. The SCS-R is an 8-item measure of perceived social belongingness (e.g., “Even around people I know, I do not feel that I really belong”) (Lee et al., 2001). Participants rated items on a 6-point Likert scale with higher scores reflecting greater social connectedness after reverse-scoring appropriate items.

The Positive and Negative Affect Scale (PANAS) is a 20-item adjective rating scale that is scored with a 5-point response format (0 – very slightly or not at all, 1 – a little, 2 – moderately, 3 – quite a bit, 4 - extremely) and grouped into general positive and negative affect sub-scales. Participants were asked to indicate the degree to which they generally feel (“that is, how you feel on the average”) the different feelings and emotions described by each adjective (Heubeck & Wilkinson, 2019; Watson et al., 1988)

The PEQ contains 145 items that evaluate life changes attributed to the dosing session and subsequent reflection. Items are rated on a 6-point Likert scale (0 = none, not at all; 5 = extreme, more than any other time in my life and more than a rating of 4). The PEQ assesses 12 subscales, including positive and negative changes in Attitudes About Life, Attitudes About Self, Mood, Social Effects, Behavioural Changes, and Spirituality (Griffiths et al., 2011). For example, pertinent Social Effects items include “You express more love toward others” and “You express more hatred toward others”. For this study, all Spirituality items were removed. All subcales were converted to percentages of their maximum values for analysis, consistent with prior work (Griffiths et al., 2018; McCulloch et al., 2022).

The Digit Symbol Substitution Test (DSST) is a computerised version of the original paper-and-pencil test taken from the Wechsler Adult Intelligence Scale (Jaeger, 2018). The participant is shown an encoding scheme consisting of a row of squares at the top of the screen, wherein nine digits are randomly associated with particular symbols. The same symbols are presented in a fixed sequence at the bottom of the screen as a row of separate response buttons. The randomisation procedure is chosen such that symbols never appear at the same ordinal position within both rows. The encoding scheme and the response buttons remain visible while the participant is shown successive presentations of a single digit at the centre of the screen. The goal is to match each digit with a symbol from the encoding list and click the corresponding response button. The number of digits correctly encoded within 3 minutes is the primary outcome. Secondary outcomes include the number of attempts, percentage accuracy (total correct/total attempts) and reaction time. Unique counterbalanced versions of the task were deployed on each test day, differing in symbol types and ordering.

The Tower of London (TOL) serves as a measure of executive functioning, specifically dimensions of mental planning and decision-making (Köstering et al., 2015). Task difficulty is dictated by elements such as the minimum number of moves required to complete the goal, goal hierarchy (i.e., ambiguity of the sequence of final moves derived from the configuration of the goal state). The present version consists of computer-generated images of beginning and end arrangements of three coloured balls. Every individual movement of the ball is counted as one step. Participants decide as quickly as possible whether the end-arrangement can be accomplished in 2, 3, 4 or 5 steps from the begin arrangement by pushing the corresponding coded button. Reaction times and total correct responses are the main dependent variables. Separate counterbalanced versions comprising unique problem sequences were provided for each test day. The TOL has been previously employed in acute studies administering ayahuasca

The Spatial Memory Test (SMT) consists of an immediate and a delayed recognition phase, serving to evaluate visuospatial memory and reasoning (Kuypers & Ramaekers, 2007). The immediate recall phase is composed of six trials in which ten black-and-white pictures (a total of 60 pictures) are subsequently presented on a computer screen at different locations for a duration of 2 s and an interstimulus interval of 1 s. After every trial, the pictures reappear one by one in the middle of the screen for 2 s, followed by the presentation of a “1” and a “2” in different locations. The participant must indicate whether each picture corresponded with either location 1 or 2. The delayed recognition phase is completed after a 30 min delay, in which the participant must indicate the correct picture location.

**S5. Methods: MRI acquisition**

A 32-receiving-channel head array Nova coil (NOVA Medical Inc., Wilmington, MA) was used to acquire imaging data for all participants on the 7T MAGNETOM scanner for an imaging session which spanned a time range of ± 75 to ± 130 min after administration. Scanning comprised structural imaging, magnetic resonance spectroscopy, task and rsfMRI.

Structural T1-weighted (T1w) images were collected at the beginning of each imaging session at an isotropic resolution of 0.9mm using a magnetisation-prepared 2 rapid acquisition gradient-echo (MP2RAGE) acquisition (190 sagittal slices following parameters: repetition time (TR) = 4,500 ms, echo time (TE) = 2.39 ms, inversion times TI1/TI2 = 900/2750 ms, flip angle1 = 5°, flip angle2 = 3°, voxel size = 0.9 mm isotropic, bandwidth = 250 Hz/pixel).

High-resolution blood oxygenation level-dependent (BOLD) signal estimation was performed using an echo planar imaging sequence of 1.5 mm isotropic voxel size (TR = 1400 ms; TE = 21 ms; field of view=198 mm; flip angle = 60°; oblique acquisition orientation; interleaved slice acquisition; 72 slices; slice thickness = 1.5 mm). Across all anatomical scans for a given subject, 7T MP2RAGE signal inhomogeneity was normalised by reconstructing "robust" T1w equivalents. In sum, as per (O'Brien et al., 2014), a normalised complexity ratio was extrapolated from T1w (GRETI1) and PDw (GRETI2) image volumes and applied to generate a uniform T1w image volume of minimal signal intensity variance.

Resting-state data (516 volumes, 12 minutes) were acquired approximately 110 minutes after substance intake. They were immediately followed by five EPI volumes in the opposite phase direction for magnetic susceptibility distortion correction using FSL *topup (Smith et al., 2004)*. During this time, participants were shown a black fixation cross on a white background and were instructed *to "Please look at the cross at all times and try to clear your mind. Stay as still as possible and do not fall asleep"* as per prior work (Mallaroni, Mason, et al., 2024; Mason et al., 2020).

**S6. Methods: Meta-analytic region of interest coordinate extraction**

Coordinates for regions of interest were identified using NeuroSynth, an automated large-scale meta-analytic platform that synthesises results from more than 14,000 published fMRI studies (https://github.com/neurosynth/neurosynth). NeuroSynth searches the neuroimaging literature for systematically reported terms (e.g., "dorsomedial prefrontal cortex") and links them to voxel-level activation coordinates mentioned in the associated papers, producing volumetric association test maps. These maps indicate the probability that a given voxel is reported in studies referencing a particular concept, without distinguishing whether the voxel is activated or deactivated, nor the magnitude of activation, but rather whether it is consistently reported in conjunction with that term. NeuroSynth is fully automated, enabling a rapid and comprehensive synthesis across the broader neuroimaging literature. For the present study, we utilised NeuroSynth association maps to identify canonical peak coordinates within five regions consistently implicated in Self–Other processing, as identified in meta-analyses, including the dorsomedial prefrontal cortex (dmPFC), ventromedial prefrontal cortex (vmPFC), precuneus (Pc), and bilateral temporoparietal junction (TPJ). Maps were averaged if multiple files contributed to a given region (eg, "cortex dmpfc", "dorsomedial prefrontal cortex", "dmpfc"). Peak voxels were then identified either across the whole brain or separately within each hemisphere, depending on the region of interest; for bilateral regions (e.g., TPJ), the peak voxel with the maximum association value was extracted independently for the left and right hemispheres. Spherical ROIs (8-mm radius) were centred on these peak coordinates and used for time series extraction.

**S7. Methods: Dynamic causal modelling (DCM)**

As we had three scanning sessions, and therefore three separate effective connectivity maps, for each participant, we created subject-specific 2nd-level effective connectivity maps by fitting a parametric empirical Bayes (PEB) model to each participant's three 1st-level DCMs. The PEB technique fits a hierarchical model to the estimated connectivity parameters and the precisions of those parameters. For this 2nd-level, the PEB model was specified with a 3x2 design matrix. The first column was a column of ones, to model baseline effective connectivity during the placebo session, and the second column was coded as ones for the 2C-B and psilocybin sessions, and a zero for the placebo session [1,1,0]. This column, therefore, modelled the difference in effective connectivity between the placebo session and the two psychedelic sessions for an individual participant.

We then adopted a PEB-of-PEBs approach to construct a 3rd-level model to make group-level inferences. Here, we fitted a single PEB model to explain the subject-specific effective connectivity maps at the 2nd level. This 3rd-level model was specified with a two-column design matrix, with a row for each participant. The first column was a column of ones. The second column was a continuous numerical variable derived from the behavioural data, specifically the Self-Other mergence score in the placebo session, subtracted from the averaged Self-Other mergence metric from the two psychedelic sessions. This variable thus represents the drug-induced change in Self-Other mergence that was captured by the behavioural model.

The 3rd-level PEB-of-PEBs model resulted in four group-level effective connectivity maps as follows: (1) Group-average baseline (placebo), (2) Group-average [drug–placebo], (3) Associations between baseline (placebo) effective connectivity and drug-induced Self-Other mergence, (4) Associations between drug-induced changes in effective connectivity and drug-induced Self-Other mergence. These maps were pruned using exploratory Bayesian model reduction and Bayesian model comparison to find the best (and simplest) model to explain the group-level data. An automatic greedy search over reduced models iteratively discarded parameters that didn't contribute to the model evidence. A Bayesian model average of parameters was then calculated over the 256 models from the final iteration of the greedy search (default). Finally, the posterior probability of each effective connectivity parameter was assessed. If the posterior probability of the parameter being non-zero was less than 0.99, the parameter was excluded from further primary analysis. For completeness, we report connections with a posterior probability of greater than 0.5 in Figure S4.

It should be noted that in DCM, the self-connections are always modelled as inhibitory (to preclude any runaway excitation), but these parameters in the model are log-scaled for the sake of numerical stability of the model fitting procedures. This (log) scaling means that these self-connections can take both positive (red) and negative values (blue). A positive self-connection means a relative increased inhibition, whereas a negative self-connection means a relative decreased inhibition (i.e., disinhibition). Inhibitory self-connections control the regions' gain or sensitivity to inputs. Only the self-connections are log-scaled in DCM.

**S8. Results: pFBT parameter estimates**

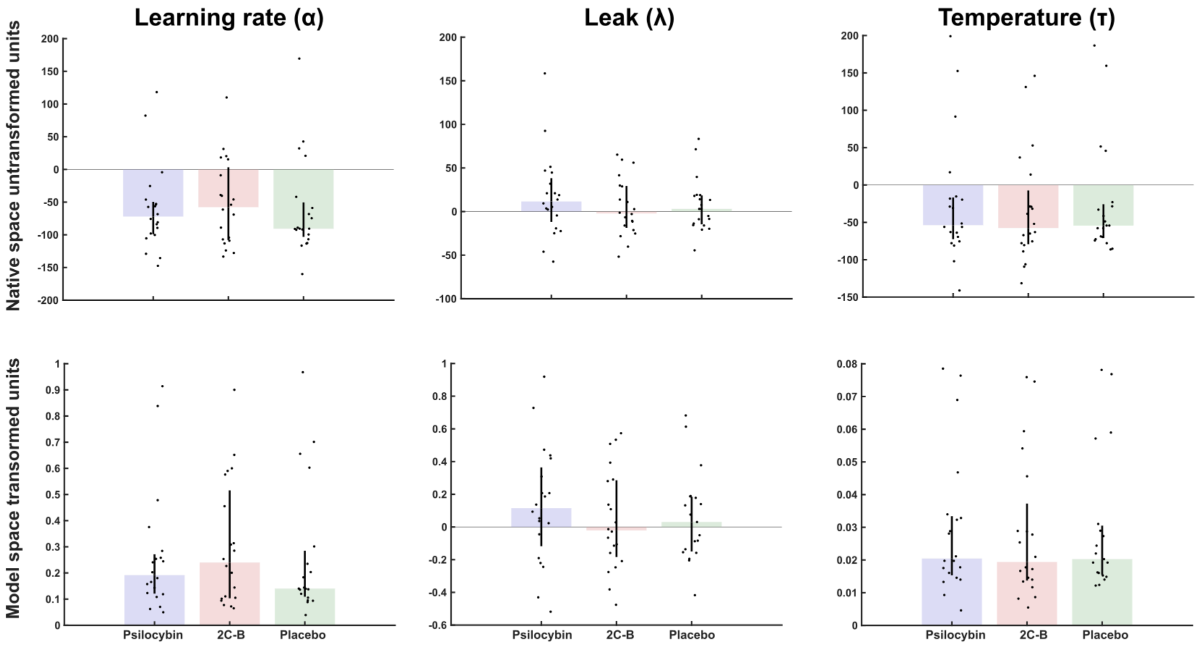


**Figure S1a. Parameter estimates.** The top row shows raw parameter values in unconstrained native space before bounded transformation. The bottom row shows parameter values in transformed model space. The bar graph shows the median +/- IQR and each dot represents a different participant.

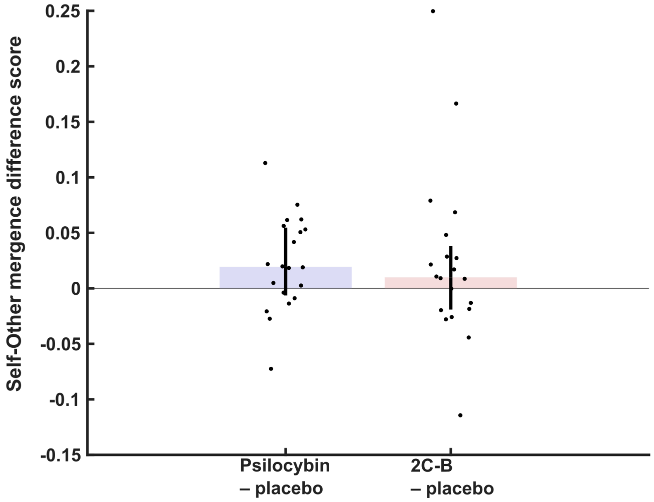


**Figure S1b. Self-Other mergence difference scores.** Difference scores showing the difference in Self-Other mergence ($\Lambda$) between psilocybin and placebo, and 2C-B and placebo. The bar graph shows the median +/- IQR and each dot represents a different participant.

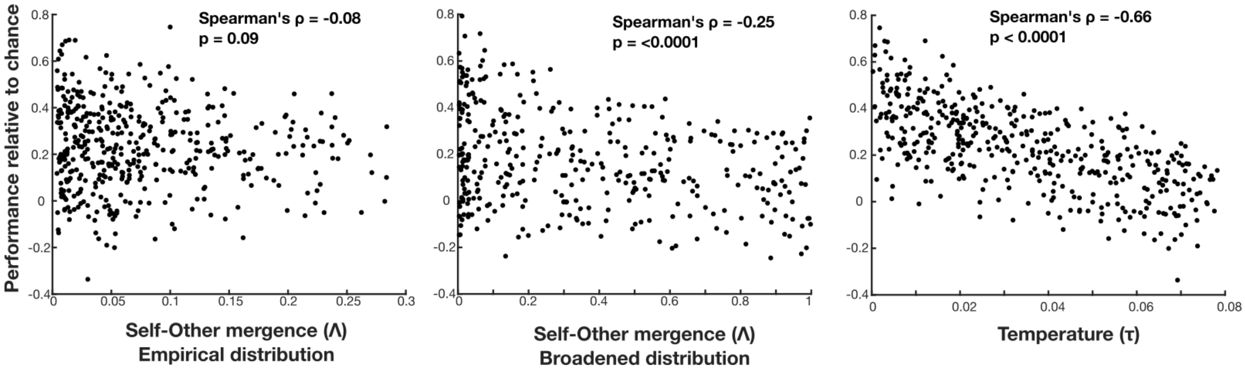


**Figure S1c. Associations between pFBT parameters and task performance.** We simulated behaviour of 500 participants by randomly sampling parameter combinations from the distribution of our empirical dataset. For each simulated subject we computed performance relative to chance using the same method as for our empirical dataset. We found that Self-Other mergence ($\Lambda$) is not associated with task performance (left). However, when simulating data using parameters from a broader distribution, which includes more extreme parameter values, we can see that Self-Other mergence ($\Lambda$) is negatively associated with task performance (middle). As expected, temperature (i.e. choice stochasticity) is negatively associated with task performance (right).

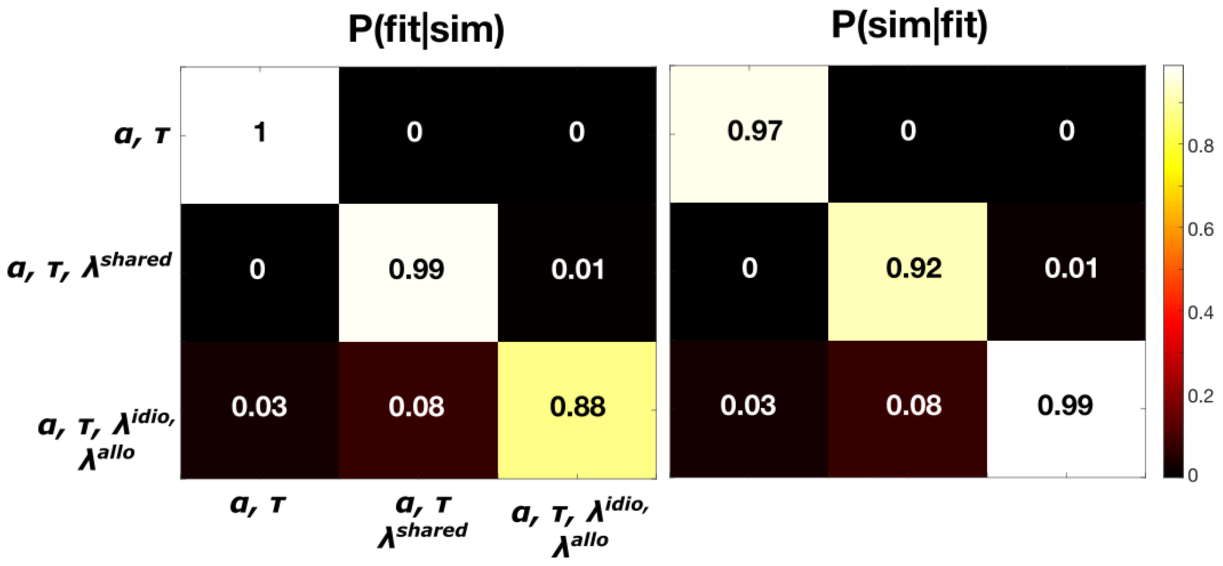


**Figure S1d. Model Recovery analysis.** To ensure we were adequately powered to run our model comparison we conducted a model recovery analysis. We chose 3 competing models. The first model was a simple 2-parameter model that included only a learning rate ($\alpha$) and temperature ( $\tau$) without any leak ($\lambda$). The second model was the same as the first model with the addition of a single leak parameter that governed both idiocentric and allocentric updating; this was the winning model in our empirical model comparison analysis. The third model was the same as the first model but with two separate leak parameters, one for idiocentric updating and one for allocentric updating. We used each of these 3 models to simulate 96 synthetic datasets, each dataset of equal sample size to our empirical dataset. We then fitted all 3 models to all datasets. We calculated the proportion of datasets for which each of the 3 models was the best-fitting model. This is visualised in the left-sided confusion matrix P(fit|sim), which shows the probability that any model was the best-fitting model, given that it was indeed the model that simulated the data. For instance, the 0.99 in the middle cell indicates that if the 3-parameter model simulated all datasets, then it is correctly identified as the winning model in 99% of those datasets. We then used Bayes rule with uniform priors (by normalising over summed columns) to compute the posterior P(sim|fit), visualised on the right. This shows the probability that a model simulated the data, given that it was the best-fitting model. For instance, the 0.92 in the middle cell indicates that if the 3-parameter model were the best-fitting model, then there is a 92% chance that it was indeed the model that generated all of the data.

#### S9. Results: Behavioural associations

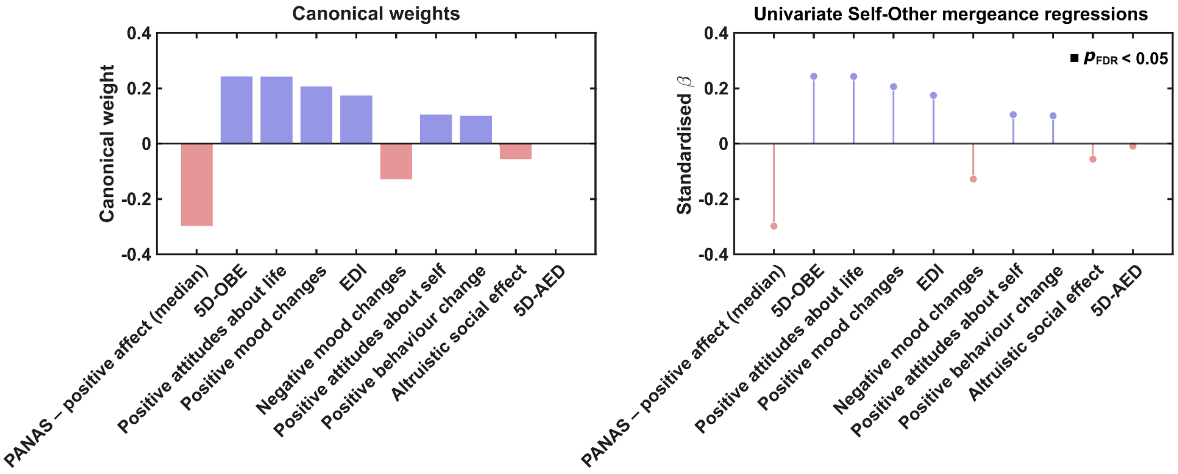


**Figure S2. Supplementary behavioural canonical variate findings.** Canonical weights are from the first canonical variate, sorted by absolute magnitude. Bars indicate the contribution of each outcome to the canonical dimension, with blue bars reflecting positive weights and red bars negative weights. Independent univariate regressions of residualised self-other mergeance scores are reported against each outcome (standardised β coefficients). Vertical stems and dots represent the sign and size of associations, coloured by direction (blue = positive, red = negative). Black squares indicate associations surviving FDR correction (*p*<0.05). Outcomes are ordered identically in both panels to enable direct comparison.

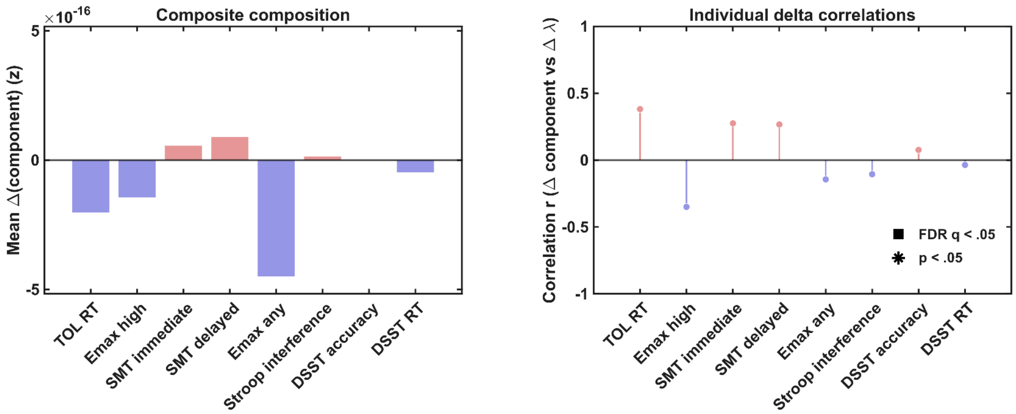


**Figure S3. Impairment burden associations.** Left, mean change (psychedelic minus placebo) for each composite component after standardisation (visualisation z), ordered by the absolute magnitude of the component–Δmergence correlation shown in the right panel. Right, (raw) Pearson correlations between each component's change and Δmergence. Vertical stems and dots represent the sign and size of associations, coloured by direction (blue = positive, red = negative). A black star denotes uncorrected $p < 0.05$, and a black square denotes FDR $q < 0.05$.

#### S10. Results: DCM quality control

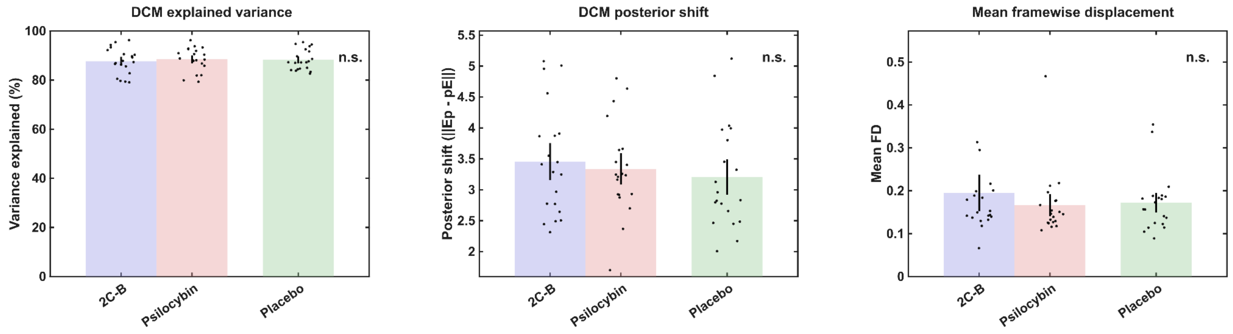


**Figure S4. Quality control measures across conditions.** Bars show mean +/- 1.5 SEM across subjects; black dots are individual subjects (N = 20). *QC metrics***:** variance explained = % variance in regional responses captured by the DCM fit (higher is better); posterior shift (||Ep - pE||) = Euclidean distance between posterior and prior parameter means across all DCM parameters, indexing how much the data moved the priors (larger = more data influence); mean FD = average framewise displacement in mm during fMRI aquisition (lower = less motion). A repeated-measures linear mixed-effects model on all subjects found no main effect of condition (any) for any metric: variance explained $F(2,38)=0.15$, $p=0.861$; posterior shift $F(2,38)=0.39$, $p=0.680$; mean FD $F(2,38)=0.69$, $p=0.506$. "n.s." marks non-significant effects.

#### S11. Results: Effective connectivity matrices

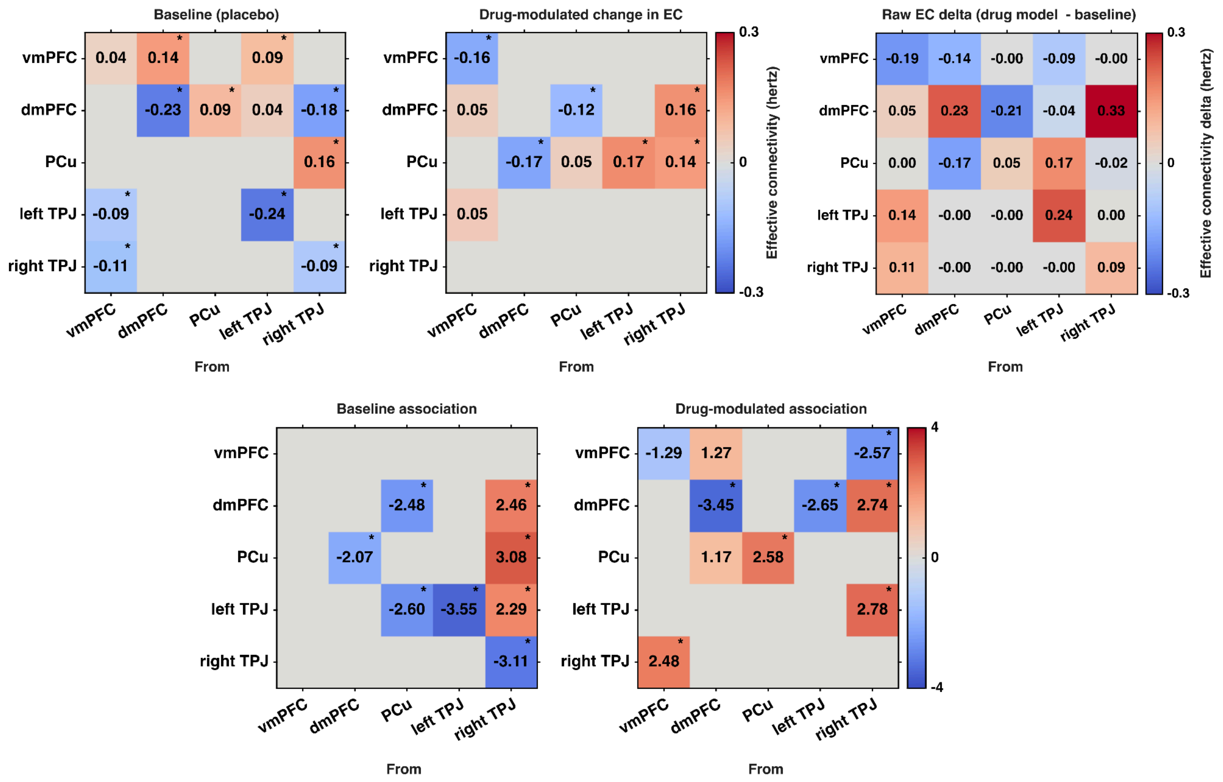


**Figure S5. Effective connectivity matrices**. **Top:** first-level DCMs for each session (placebo, psilocybin, 2C-B) were combined into participant-level models estimating baseline EC (placebo) and drug-induced change ([drug – placebo]). **Bottom:** a third-level PEB-of-PEBs tested group-average effects and associations with Self–Other mergence. At baseline, blue denotes inhibitory and red excitatory influences (Hz; self-connections log-scaled and inherently inhibitory). For drug–placebo contrasts, arrows indicate increases (red) or decreases (blue) relative to baseline, not absolute valence; this distinction matters as some connections may flip sign under drug. Association strength is reported in arbitrary units, following the same visual convention as the EC strength results. Connections shown in the main text meet posterior probability >0.99 (★), with weaker evidence (≥0.5) shown here. Abbreviations: dmPFC = dorsomedial prefrontal cortex; vmPFC = ventromedial prefrontal cortex; PCu = precuneus; lTPJ = left temporoparietal junction; rTPJ = right temporoparietal junction.

#### S12. Results: Exploratory DCM multivariate associations

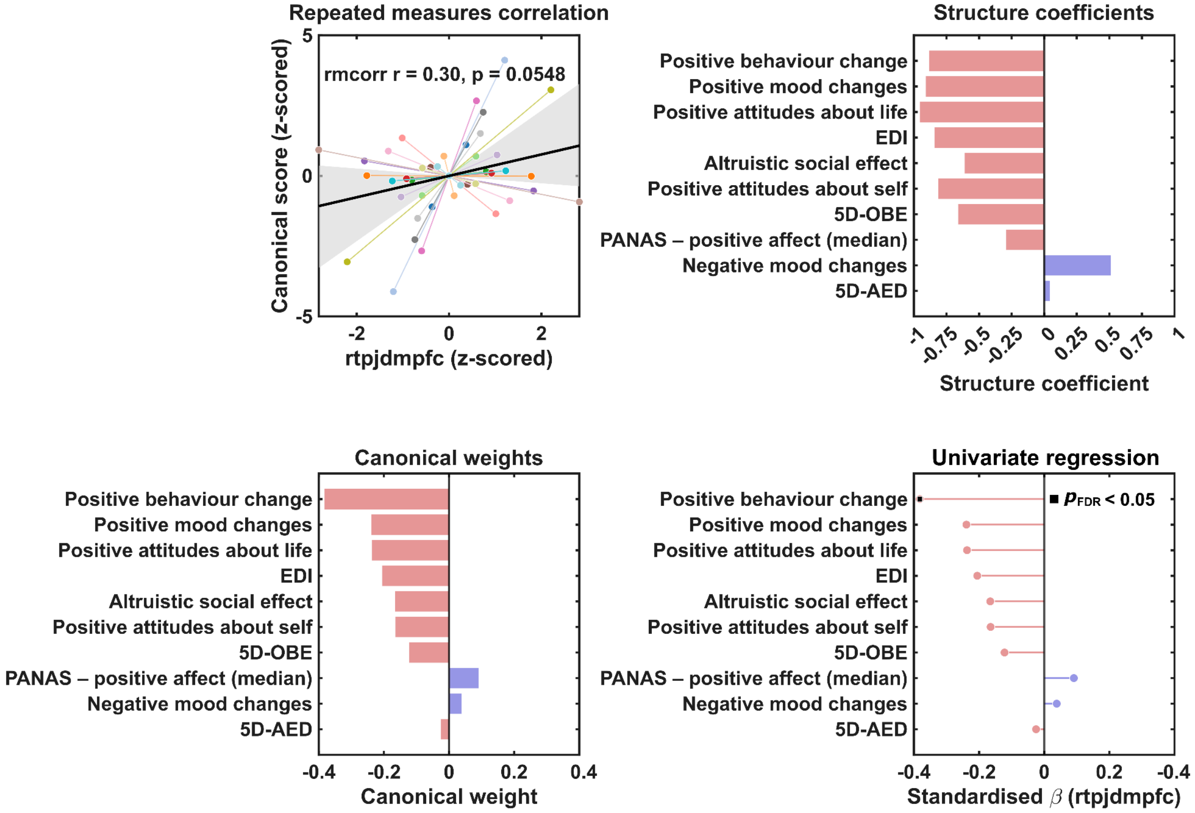


**Figure S6. Exploratory multivariate association between rTPJ→dmPFC effective connectivity and subjective outcomes.** Exploratory analyses suggested that attenuation of the inhibitory rTPJ→dmPFC connection under psychedelics was related to psychosocial outcomes. A within-subject permutation MANOVA indicated a trend-level multivariate association (Pillai's trace = 0.33, $p_{perm}$ = 0.063). Canonical analysis revealed a significant correlation between the connection and the first canonical variate (r = 0.30, $p_{perm}$ = 0.017), with a repeated-measures correlation showing a trend toward a within-participant effect (r = 0.30, $p_{within}$ = 0.055). However, cross-validation did not support out-of-sample generalisation (r = 0.12, $p_{perm}$ = 0.65), suggesting that these effects should be interpreted cautiously. Negative coefficients indicate that participants showing a stronger loss of inhibition in the rTPJ→dmPFC connection also showed greater positive psychosocial effects; conversely, more inhibitory connectivity was associated with fewer positive effects. Repeated-measures scatter showing the association between rTPJ-> dmPFC effective connectivity and the canonical (sub)acute psychosocial response score. Points are drug-aware, subject-level residuals (z-scored); line segments link sessions from the same participant. The black line is the common repeated-measures correlation fit; the shaded band is its 95% CI from a subject-level bootstrap. Because residualisation includes an intercept and drug term, placebo observations cluster near the origin. Structure coefficients (correlation of each outcome with the canonical variate) indicate how strongly each measure aligns with the multivariate association. ts. Canonical weights from the first canonical variate, ordered by absolute magnitude. Bar colour encodes direction (blue = positive, red = negative). Univariate regressions of residualised rTPJ→dMPFC effective connectivity on each outcome (standardised β). Vertical stems and dots show effect size and sign (blue = positive, red = negative); black squares mark effects surviving FDR correction ($p < 0.05$). Outcomes are ordered identically to enable direct comparison.

**S13. Results: DCM specificity analyses**

Recent computational accounts argue that what is often labelled “social cognition” is better framed in terms of the computations a behaviour requires, with the same regions being repurposed across social and non-social settings when the underlying demands are similar. In this view, the rTPJ may support predictive and inference-like operations that can generalise across contexts, and apparent differences between social and non-social conditions could partly reflect behavioural relevance and attention rather than a uniquely social mechanism (Mahmoodi & Rushworth, 2026; Wilterson et al., 2021).

Given this perspective, we tested whether the pFBT self-other boundary phenotype could be alternatively explained by the TPJ being part of a non-ToM task-active network (see Figure S7a). Following the same meta-analytic approach as before, we specified another 5-node TPJ salience/ventral-attentional network model consistently implicated in psychedelic action and the triple network theory of psychopathology (Doss et al., 2022; Menon, 2019), comprising the same bilateral TPJ coordinates (left: [-50.0, -56.0, 22.0]; right: [58.0, -56.0, 18.0]); bilateral anterior insula (left: [-36.0, 16.0, 2.0]; right: [38.0, 20.0, 2.0]), and the dorsal anterior cingulate ([6.0, 28.0, 24.0]). We repeated the identical spectral DCM and PEB workflow used in our main analysis.

Using the same conjunction criteria as the primary results (baseline coupling, behavioural association, group-level drug effect, and drug-induced coupling changes tracking behaviour), the comparator network showed robust drug-related effects yet did not reproduce the rTPJ-centred behaviour-linked modulation pattern (Figure S7b). Neither the rTPJ nor any included region’s afferent or efferent connections met this criterion, suggesting the specificity of our main finding.

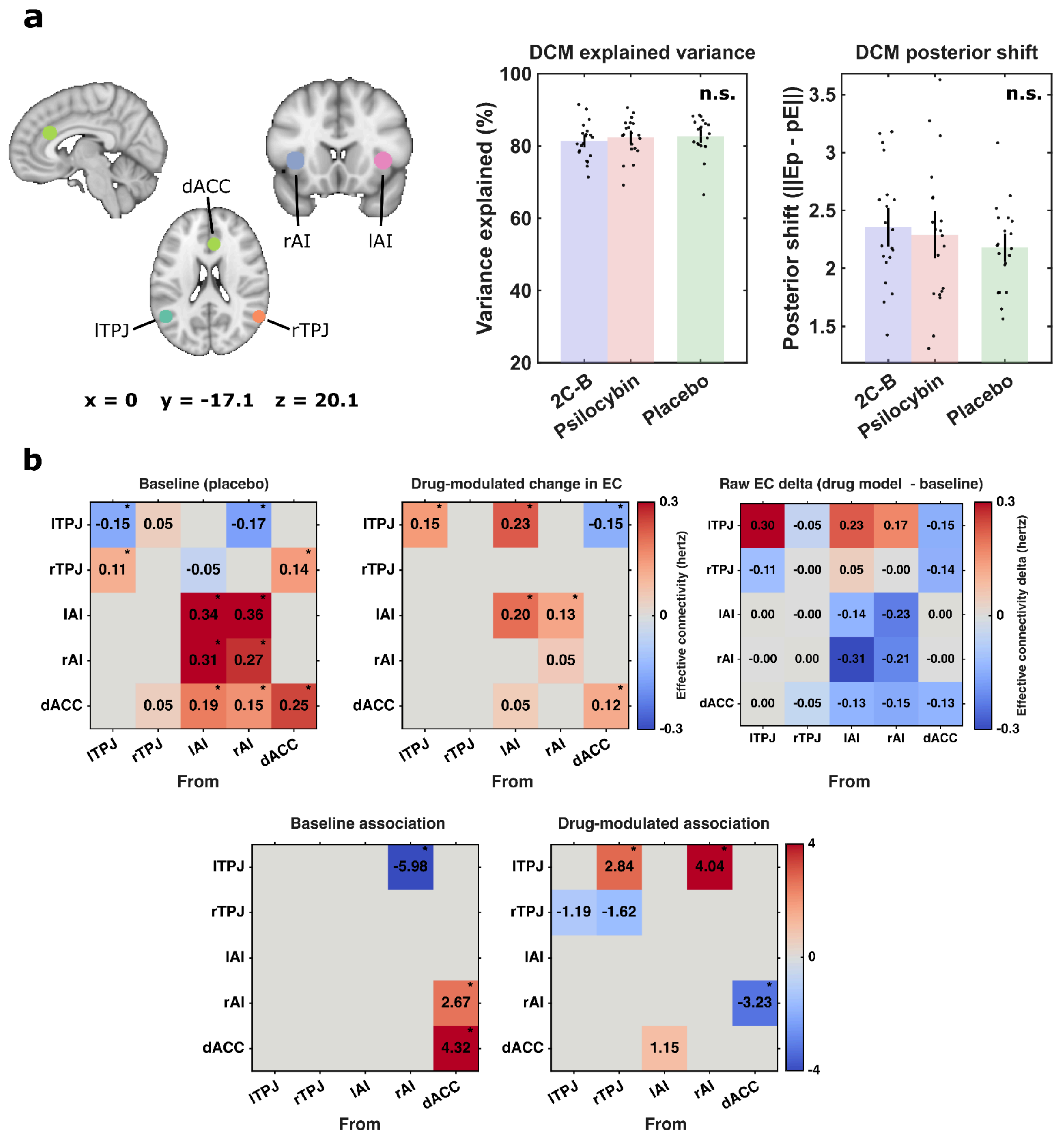


**Figure S7. Specificity of rTPJ findings using a salience comparator network.** a, Regions of interest are shown on canonical brain slices with corresponding MNI coordinates (mm). Quality control barplots show mean +/- 1.5 SEM across subjects; black dots are individual subjects (N = 20). *QC metrics*: variance explained = % variance in regional responses captured by the DCM fit (higher is better); posterior shift (||Ep - pE||) = Euclidean distance between posterior and prior parameter means across all DCM parameters, indexing how much the data moved the priors (larger = more data influence). Repeated-measures linear mixed-effects models found no main effect of condition for any metric: variance explained $F(2, 38) = 0.34$, $p = 0.7157$; posterior shift

$F(2, 38) = 0.08$, $p = 0.9211$. “n.s.” marks non-significant effects. DCM specificity analyses. Effective connectivity analyses here were performed identically to our primary approach, embedding instead using TPJ coordinates in a salience-ventral attentional network. **Top:** First-level DCMs for each session (placebo, psilocybin, 2C-B) were combined into participant-level models estimating baseline EC (placebo) and drug-induced change ([drug – placebo]). **Bottom:** a third-level PEB-of-PEBs tested group-average effects and associations with Self–Other mergence. At baseline, blue denotes inhibitory and red excitatory influences (Hz; self-connections log-scaled and inherently inhibitory). For drug–placebo contrasts, arrows indicate increases (red) or decreases (blue) relative to baseline, not absolute valence; this distinction matters as some connections may flip sign. Association strength is reported in arbitrary units, following the same visual convention as the EC strength results. Connections shown in the main text meet posterior probability >0.99 (★, none), with weaker evidence (≥0.5) shown here.
